# Disease-specific prioritization of non-coding GWAS variants based on chromatin accessibility

**DOI:** 10.1101/2023.10.17.23297164

**Authors:** Qianqian Liang, Abin Abraham, John A Capra, Dennis Kostka

**Affiliations:** Department of Developmental Biology, University of Pittsburgh School of Medicine, Pittsburgh, PA, USA; Department of Human Genetics, University of Pittsburgh School of Public Health, Pittsburgh, PA, USA; Children’s Hospital of Philadelphia, Philadelphia, PA, USA; Department of Epidemiology & Biostatistics and Bakar Computational Health Sciences Institute, University of California, San Francisco, CA, USA; Department of Computational & Systems Biology and Center for Evolutionary Biology and Medicine, University of Pittsburgh School of Medicine, Pittsburgh, PA, USA

## Abstract

Non-protein-coding genetic variants are a major driver of the genetic risk for human disease; however, identifying which non-coding variants contribute to which diseases, and their mechanisms, remains challenging. In-silico variant prioritization methods quantify a variant’s severity in the context of having a phenotypic effect; but for most methods the specific phenotype and disease context of the prediction are poorly defined. For example, many commonly used methods provide a single organism-wide score for each variant, while other methods summarize a variant’s impact specifically in certain tissues and/or cell-types. Here we propose a complementary disease-specific variant prioritization scheme, which is motivated by the observation that the variants contributing to different diseases often operate through different biological mechanisms.

We combine tissue/cell-type specific scores into disease-specific scores with a logistic regression approach and apply it to 25,000 non-coding variants spanning 111 diseases. We show that disease-specific aggregation of tissue/cell-type specific scores (GenoSkyline, Fit-Cons2, DNA accessibility) signifiantly improves the association of common non-coding genetic variants with disease (average precision: 0.151, baseline=0.09), compared with organism-wide scores (GenoCanyon, LINSIGHT, GWAVA, eigen, CADD; average precision: 0.129, base-line=0.09). Calculating disease similarities based on data-driven aggregation weights highlights meaningful disease groups (e.g., immune system related diseases and mental/behavioral disorders), and it provides information about tissues and cell-types that drive these similarities (e.g., lymphoblastoid T-cells for immune-system diseases). We also show that so-learned similarities are complementary to genetic similarities as quantified by genetic correlation. Overall, our aggregation approach demonstrates the strengths of disease-specific variant prioritization, leads to improvement in non-coding variant prioritization, and it enables interpretable models that link variants to disease via specific tissues and/or cell-types.

## 1 Introduction

Characterizing non-coding genetic variants in the human genome is essential for making progress toward better understanding the genetic components of disease, because ∼90% of disease-associated variants discovered by genome-wide association studies (GWAS) are located in non-protein-coding regions [1]. Further on, whole-genome sequencing (WGS) discovers disease-associated variants genome-wide [2, 3] and is increasingly becoming an assay of choice. Therefore, approaches for characterizing and prioritizing non-coding variants can be expected to play an increasingly important role, especially when assessing discovered variants in the context of functional follow-up experimental studies.

Efforts to computationally characterize and better understand non-coding variants take advantage of sequence, functional genomics, comparative genomics, and epigenomics data [4, 5, 6], and more. These data are combined and used to train and develop supervised and/or unsupervised models that attempt to quantify a variant’s impact [7]. We find it conceptually useful to distinguish between variant scores that model overall impact (that is on the level of the whole organism, *orgnaism-level* scores) and scores that quantify impact in a specific context, like a tissue or a cell-type (i.e., *tissue-level* scores). Examples for organism-level scores are CADD [8], Eigen [9], or LINSIGHT [10], while scores from methods like GenoSkyline [11], Fitcons2 [12], or FUN-LDA [13] are tissue-specific.

Often interest in a set of variants is from the perspective of studying a specific disease. In that case, organism-level scores are likely to be overly general. That is, a variant’s impact might be considered high because it disrupts the functional role of a sequence element. However, that functional role may be unrelated to the disease of interest. In one study, for instance, organism-level scores like CADD and DANN were unable to discover an enrichment signal for brain-related traits, while context-specific variant scores focusing on relevant tissues were successful [14]. This demonstrates that tissue-specific scores can address the issue of disease specificity to some extent. However, aspects of disease-relevant tissues typically remain unknown, and often more than one tissue is implicated with a specific trait (termed “multifactorial” and “polyfactorial” traits) [15]. This suggests the use of *disease-specific* variant scores that characterize variants in the context of a specific disease phenotype of interest.

Computational methods for disease-specific variant prioritization do exist. Some approaches are geared towards one disease (e.g, congenital heart disease [16], amyotrophic lateral sclerosis [17]) or towards a specific class of diseases (e.g., autoimmune diseases [18]). This focus prevents them from being readily adapted to other disease types. Others, like DIVAN [19], PINES [20], and ARVIN [21], cover a broader range of disease types. Of these, ARVIN requires a priori knowledge of disease-relevant tissues, whereas DIVAN and PINES do not. PINES uses an enrichment-based method to predict and up-weight disease-relevant tissues/cell-types, whereas DIVAN uses a more complex machine learning algorithm. The PINES approach has been evaluated on a relatively small set of traits (∼10 different contexts), while DIVAN’s more compex model renders understanding the relationship between different tissues and diseases difficult.

In this work, we derive disease-specific variant scores by combining published tissue-specific scores. We use a carefully regularized logistic regression approach to derive data-driven disease-specific combination weights, which allow us to better associate variants with disease. In addition, they enable us to quantify a similarity between different disease phenotypes. Using the NHGRI-EBI GWAS catalog [1] we compiled a benchmark dataset containing about 63k phenotype-associated non-protein-coding single nucleotide variants across 111 disease phenotypes (together with matched random controls). We then demonstrate that using disease-specific combination weights outperforms conventional organism-level approaches, that our interpretable model has competitive performance, and that it enables a disease similarity measure that captures information complementary to established measures like genetic correlation.

## 2 Results

### 2.1 Non-coding GWAS variants associated with disease phenotypes, and matched controls

In order to study variant prioritization methods, we created a dataset of “positive” (i.e., disease associated) non-coding variants, matched with a random set of “negative” or “control” variants. This setup allowed us to quantitatively assess prioritization methods based on their performance in discriminating positive from control variants.

#### 2.1.1 Disease-associated non-coding SNVs

We used a subset of single nucleotide variants (SNVs) reported in the EBI/NIH GWAS catalog [1] to compile an inventory of disease-associated non-coding variants. Specifically, we focused in reported variants that *(a)* do not overlap protein-coding sequence (see **Methods**) and *(b)* that are associated with a disease phenotype as noted in the Experimental Factor Ontology (EFO) trait description, which is provided within the catalog. We define disease phenotypes as descendants of the EFO term “disease” (EFO:0000408). Focusing on disease terms with at least 100 annotated SNVs resulted in 26,080 associations involving 20,656 SNVs and 67 disease phenotypes. The EFO provides parent-child relations between disease terms (parent = more general, child = more specific), and propagating SNVs from child-terms to parent-terms increased the number of disease phenotypes with at least 100 SNVs, resulting in 77,028 association between 25,516 SNVs and 111 diseases. We find that most of the SNVs we recover are located in intronic (60.5%) and intergenic (25.8%) sequence (**Fig. 1A**), and that a majority of SNVs are directly annotated to a single disease phenotype (**Fig. 1B**). After propagating annotated SNVs from child to parent terms, SNV-to-disease annotations become predominantly many:many (**Fig. 1B**). **Suppl. Data** SD1 lists disease terms and corresponding numbers of disease-associated SNVs.

**Figure 1.**
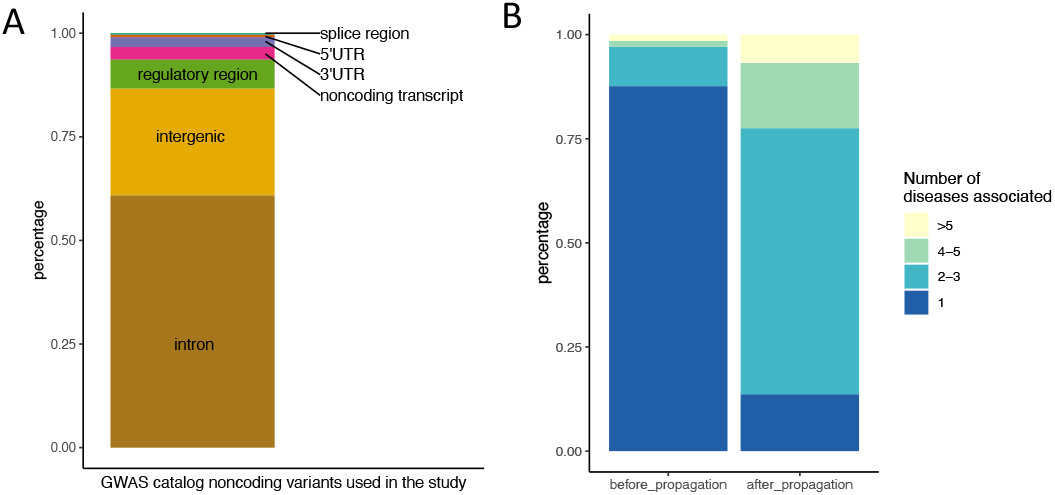
Disease-associated non-coding SNVs. (A) Genomic context of non-coding SNVs used in this study. (B) Percentage of the SNVs used that are annotated to 1, 2-3, 4-5 or more than 5 disease phenotypes, before and after propagating SNV-phenotype associations according to EFO parent-child annotations. Genomic context annotation is adapted from the CONTEXT column from the GWAS catalog, where we combine splice donor, splice region and splice acceptor variants into splice variants and I combine TF binding variants and regulatory regions variants into regulatory region variants.

#### 2.1.2 Control SNVs

For each disease-associated SNV we selected ∼10 matched control-SNVs using a re-implementation of the SNPsnap approach [22], while avoiding duplicate control-SNV across the overall dataset (see **Methods**). This yielded 255,137 control SNVs (for some disease associated SNVs we could not retrieve the full ten control SNVs). With these results we have access to data for 111 disease terms, containing disease-associated SNVs together with matched controls. **Suppl. Data** SD2 and SD3 contain information about all disease and control SNVs used in this study, respectively.

### 2.2 Disease-specific non-coding variant prioritization with organism-level variant scores is only moderately successful

We assessed how well current commonly-used organism-level variant scores are able to prioritize disease-associated vs. control-SNVs for the 111 disease terms we studied. **Fig. 2** summarizes results, where boxplots of two performance measures (area under the ROC curve and average precision (= area under the precision recall curve)) are shown for CADD [8], eigen [9], GenoCanyon [11], GWAVA [23], and LINSIGHT [10] scores. We find that organism-level scores, while improving upon random guessing, are only moderately successful in correctly prioritizing disease-associated non-coding variants. Comparing variant scores with each other we find that relative performance differences appear overall robust with respect to the metric employed (area under the ROC curve vs. average precision). It is qualitatively visible that CADD performs less favorably than other methods, but also that there are differences between these. We therefore compared performance between different scores in more detail.

**Figure 2.**
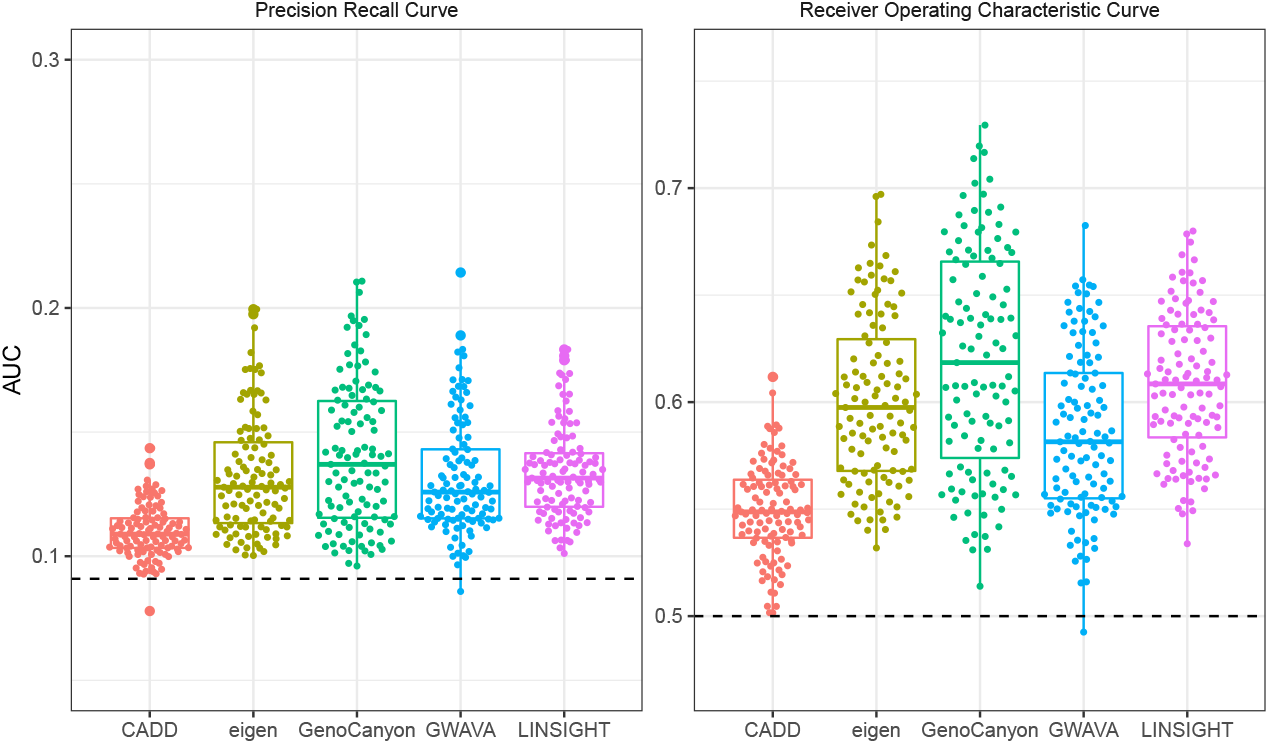
Organism-level variant scores are moderately successful in prioritizing non-coding disease-associated variants. Different organism-level variant prioritization scores are shown on the x-axis, the y-axis displays performance in terms of average precision (area under the precision recall curve, left panel) and area under the receiver-operator curve (right panel). Each point represents a specific disease term from the experimental factor ontology. Horizontal lines spanning data sets show expectations under random guessing.

We studied the performance of different scores at two levels of resolution: In aggregate across all disease terms, and for each disease term separately. For both approaches we used Wilcoxon signed-ranks tests to decide whether one score significantly outperforms another score (= significant p-value) or whether performance is tied (= non-significant p-value); see **Methods** section. Results are summarized in **Tab**. 1. We find that GenoCanyon has better performance compared with other variant scores, followed by LINSIGHT, GWAVA and eigen, while CADD is consistently outperformed by other methods. Performance differences between LINSIGHT, GWAVA and eigen are not significant when aggregating across disease terms (last three columns in **Tab**.1); however, when counting individual terms LINSIGHT has most wins and fewest losses, while eigen has most losses and fewest wins, leading to the ordering displayed in **Tab**.1. **Suppl. Data** SD4 and SD5 contain results for all comparisons. Overall these quantitative results are in-line with the visual impression from **Fig**. 2. Next, we investigated if the performance of organism-level variant scores could be improved by using tissue-specific scoring approaches.

**Table 1:**
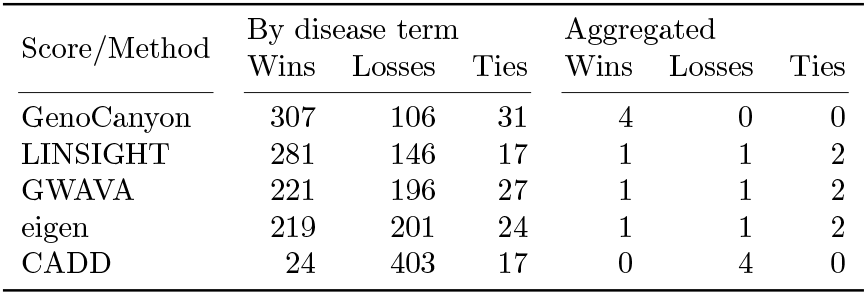
Relative performance of organism-level variant scores. Wins, Losses, Ties refers to significantly better (or worse, or tied) performance across all possible pairings (see **Methods**). The first three columns summarize separate comparisons for each disease term (for each row there are four other methods and 111 terms, i.e. 444 comparisons), while the last three columns represent results of comparisons between scores aggregated across terms. Average precision was used as the performance metric, and Wilcoxon singed-ranks tests to determine wins and losses (p-values less than 0.05 are reported as ties).

| Score/Method | By disease term |  |  | Aggregated |  |  |
| --- | --- | --- | --- | --- | --- | --- |
|  | Wins | Losses | Ties | Wins | Losses | Ties |
| GenoCanyon | 307 | 106 | 31 | 4 | 0 | 0 |
| LINSIGHT | 281 | 146 | 17 | 1 | 1 | 2 |
| GWAVA | 221 | 196 | 27 | 1 | 1 | 2 |
| eigen | 219 | 201 | 24 | 1 | 1 | 2 |
| CADD | 24 | 403 | 17 | 0 | 4 | 0 |

### 2.3 Disease-specific scores improve non-coding variant prioritization

#### 2.3.1 Disease-specific aggregation weights for tissue-specific variant scores

We studied three tissue-specific scores for variant prioritization to explore if their usage can improve the performance of organism-level scores. Specifically, we used Genoskyline [11] and Fitcons2 [12] as scores designed to prioritize variants, and we also evaluated DNase I hypersensitivity (DHS) profiles from the ENCODE project [6]. All of these scores are available for 127 contexts [5] spanning a diverse set of cell and tissue types, including heart, brain, immune cells, and more.

For each tissue-specific score we assess two approaches to prioritize variants. First, as a baseline approach we aggregate scores across tissues in a *disease-agnostic* way. That is, for a specific variant we average scores at the variant position across all tissues (termed ***tissue-mean***), essentially producing a organism-level type score, independent of the disease term under consideration. Second, we aggregate scores across tissues in a *disease-specific* way. Briefly, we train a regularized logistic regression model for each disease term that learns disease-specific tissue aggregation weights. In a nested cross-validation setup learned weights are then applied to held-out variants, allowing for a fair performance assessment of this approach (termed ***tissue-weighted***), see **Methods. Fig. 3** summarizes our findings.

**Figure 3.**
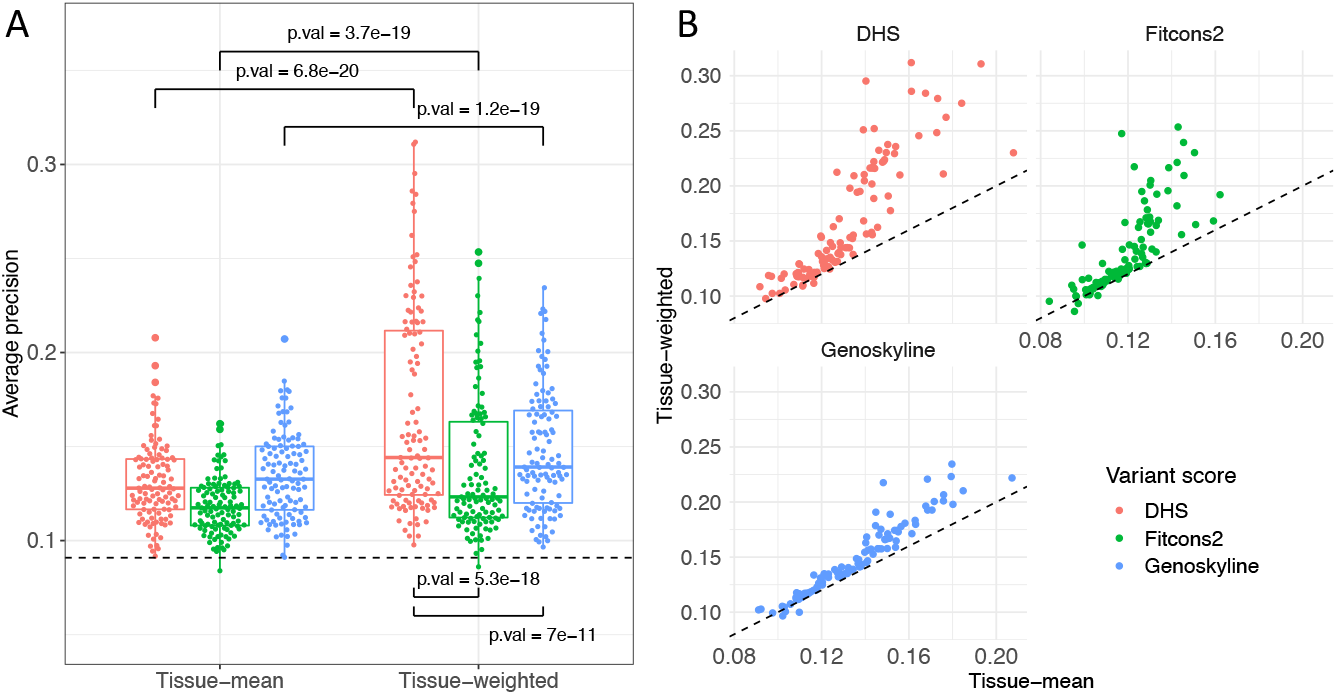
Disease-specific tissue weights improve variant prioritization. Performance of three tissue-specific variant scores (DHS, Fitcons2, Genoskyline) is used to prioritize non-coding disease-associated variants for disease terms using two approaches: *tissue-mean* (i.e., disease-agnostic, baseline) on the left side and and *tissue-weighted* (i.e., disease specific) on the right side. P-values were calculated using a Wilcoxon signed-ranks test (A). Scatter plot of tissue-mean vs. tissue-weighted performance (average precision) for each tissue-specific score; dashed line denotes the diagnonal (B).

In **Fig. 3A** we show tissue-mean performance (measured by average precision) for the three scores we study on the left, and tissue-weighted performance on the right. For all three scores tissue-weighted significantly outperforms tissue-mean (Wilcoxon signed-ranks test, p-values *<* 0.0001). **Fig. 3B** shows tissue-mean vs. tissue-weighted comparisons for each score, and we see that in almost all disease terms tissue-weighted outperforms tissue-mean. See **Suppl. Data** SD6 and SD7 for tissue-mean vs. tissue-weighted performances for each disease term, and for aggregated performances across all disease terms. The improvement remains evident if we limit disease-associated SNVs to one variant per LD block, and also when we insure that the SNVs in the training and test datasets are not on the same chromosome (See **Suppl. Fig**. S17 -S20 and the Supplemental material for more details).

While the performance-gain for tissue-weighted is broadly consistent across diseases, for some it is more pronounced than for others. To illustrate this observation, we selected four disease terms with a high performance gain, four terms with a medium gain, and four terms where we observed the least gain (Best improvement, ranking 1-4; middle improvement, ranking 20-23; least improvement, ranking 108-111). **Fig**. 4 shows our findings, where variability in tissue-weighted performance induced by varying train-test-fold splits during cross-validation is also displayed. We see that for Celiac Disease (EFO:0001060), Systemic Scleroderma (EFO:0000717), Chronic Lymphocytic Leukemia (EFO:0000095) and Sclerosing Cholangitis (EFO:0004268) performance is consistently improved for all three tissue-weighted scores, while for Retinopathy (EFO:0003839), Endometriosis (EFO:0001065), Diabetic Nephopathy (EFO:0000401) and HIV-1 Infection(EFO:0000180) we find no improvement. We also note that disease terms with pronounced improvement appear to have better baseline (i.e., tissue-mean) performance than disease terms where we find little or no benefit of the tissue-weighted approach. Improvement for diseases shown in **Fig**. 4 is largest for DHS, but, consistent with **Fig**. 3, we see improvement for Fitcons2 and Genoskyline as well.

**Figure 4.**
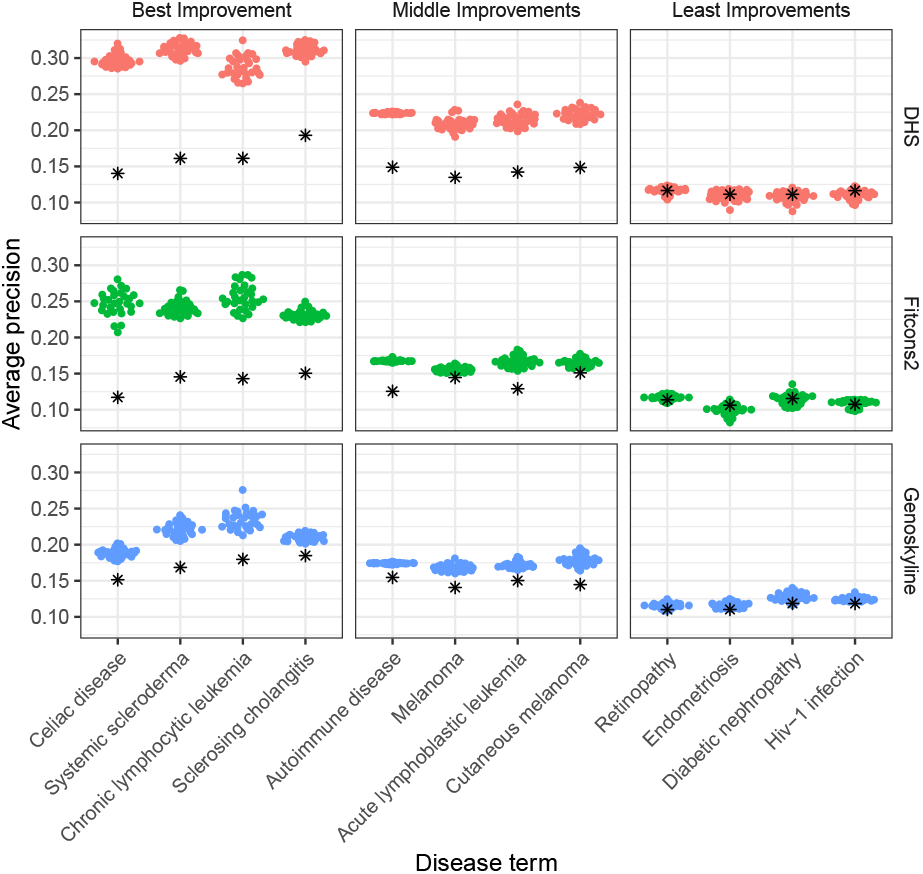
Improvement through disease-specific tissue weights is consistent across scores but varies with disease term. Shown is the performance of tissue-weighted variant scores (colored points) vs. tissue-mean (black asterisks) as a baseline, for three tissue scores (rows) and four diseases, stratified by improvement observed: best improvement for the fist column, moderate improvement for the middle column, and least improvement for the right column. X-axes denote disease terms, the y-axis average precision. Different points for tissue-weighted scores represent different data-splits in the nested cross validation procedure.

### 2.3.2 DNase I hypersensitivity (DHS) scoring outperforms other tissue specific scores

To quantify relative performance of the three different tissue-specific scores, we proceed similarly to organism-level scores. Focusing on pairwise comparisons we find that DHS scores outperform Genoskyline and Fitcons2 for most disease terms, and on average (see **Tab**. 2). This observation is consistent with **Fig**. 3 and 4, which often show higher average precision values for DHS than for the other two scores. Notably, baseline (i.e., tissue-mean) performance of DHS does not appear significantly better than that of Genoskyline (**Fig**. 3). **Suppl. Data** SD8 and SD9 contain details for comparisons between DHS, Fitcons2 and Genoskyline for all disease terms. Next, we explored whether disease-specific tissue weights outperform organism-level scores.

**Table 2:** DHS outperforms other tissue-specific scores. Wins, Losses, Ties refer to significantly better (or worse, or tied) performance across all possible score pairings (see **Methods**). The first three columns summarize separate comparisons for each disease term (for each row there are two other methods and 111 terms, i.e., 222 comparisons), while the last three columns represent results of comparisons aggregated over disease terms. Average precision was used as the performance metric, and the Wilcoxon singed-ranks test to determine wins and losses (p-values less than 0.05 are reported as ties).

| Score/Method | By disease term |  |  | Aggregated |  |  |
| --- | --- | --- | --- | --- | --- | --- |
|  | Wins | Losses | Ties | Wins | Losses | Ties |
| DHS | 180 | 22 | 20 | 2 | 0 | 0 |
| Genoskyline | 96 | 94 | 32 | 1 | 1 | 0 |
| Fitcons2 | 19 | 179 | 24 | 0 | 2 | 0 |

**Table 3:** DHS outperforms organism-level variant scores. Wins, Losses, Ties refer to significantly better (or worse, or tied) performance across all possible score pairings (see **Methods**). The first three columns summarize separate comparisons for each disease term (for each row there are two other methods and 111 terms, i.e., 555 comparisons), while the last three columns represent results of comparisons aggregated over terms. Average precision was used as the performance metric, and the Wilcoxon singed-ranks test to determine wins and losses (p-values less than 0.05 were reported as ties).

| Score/Method | By disease term |  |  | Aggregated |  |  |
| --- | --- | --- | --- | --- | --- | --- |
|  | Wins | Losses | Ties | Wins | Losses | Ties |
| DHS | 474 | 44 | 37 | 5 | 0 | 0 |
| GenoCanyon | 314 | 198 | 43 | 4 | 1 | 0 |
| LINSIGHT | 298 | 230 | 27 | 1 | 2 | 2 |
| GWAVA | 233 | 289 | 33 | 1 | 2 | 2 |
| eigen | 223 | 299 | 33 | 1 | 2 | 2 |
| CADD | 28 | 510 | 17 | 0 | 5 | 0 |

#### 2.3.3 DNase I hypersensitivity (DHS) tissue-weighted scoring outperforms organism-level variant scores

To compare the DHS tissue-weighted score with organism-level scores, we directly contrasted their performance. Similar to before, **Tab**. 3 summarizes DHS “wins” (= significantly better performance of DHS tissue-weighted, p-value ≤ 0.05), losses, and ties, compared with five organism-level variant scores, individually (i.e., per disease term) and aggregated across disease terms. In addition, **Tab**. ST4 summarizes pair-wise comparisons between tissue-weighted DHS and each organism-level score. We find that DHS tissue-weighted outperforms all organism-level scores in the aggregated analyses, and that it outperforms all other scores on the majority of disease terms (it only performs significantly worse than any other score in 44 out of 550 comparisons).

GenoCanyon is the most competitive organism-level score, where DHS is significantly better for 92 terms out of 111 (∼83%). Interestingly, LINSIGHT performs better against DHS than GenoCanyon, which is the best overall performing organism-level score (see **Tab**. ST4). **Suppl. Data** SD10 contains detailed results for each comparison. We also find that DHS outperforms organism-level scores when aggregating over disease terms (also see **Suppl. Data** SD11).

To illustrate the gain in performance, we selected four example disease terms where disease-specific variant prioritization yielded high improvements, medium improvements, comparable performance, and worse performance, respectively. Selection was based on ranking differences between DHS and GenoCanyon: best improvement, ranks 1-4; medium improvements, ranks 25-28; comparable performance, ranks 64-67; GenoCanyon better, ranks 108-111. Results are summarized in **Fig**. 5, where we find substantial improvements using tissue-weightes scoring for Systemic Sceleroderma (EFO:0000717), Celiac Disease (EFO:0001060), Sclerosing Chalangitis (EFO:0004268) and Multiple Sclerosis (EFO:0003885), for which we have already noticed substantial improvement of DHS tissue-weighted over DHS tissue-mean. Disease terms where GenoCenyon is performing better include Venous Thromboembolism (EFO:0004286), Diverticular Disease (EFO:0009959), Non-small Cell Lung Carcinoma (EFO:0003060), and Lung Adenocarcinoma (EFO:0000571).

**Figure 5.**
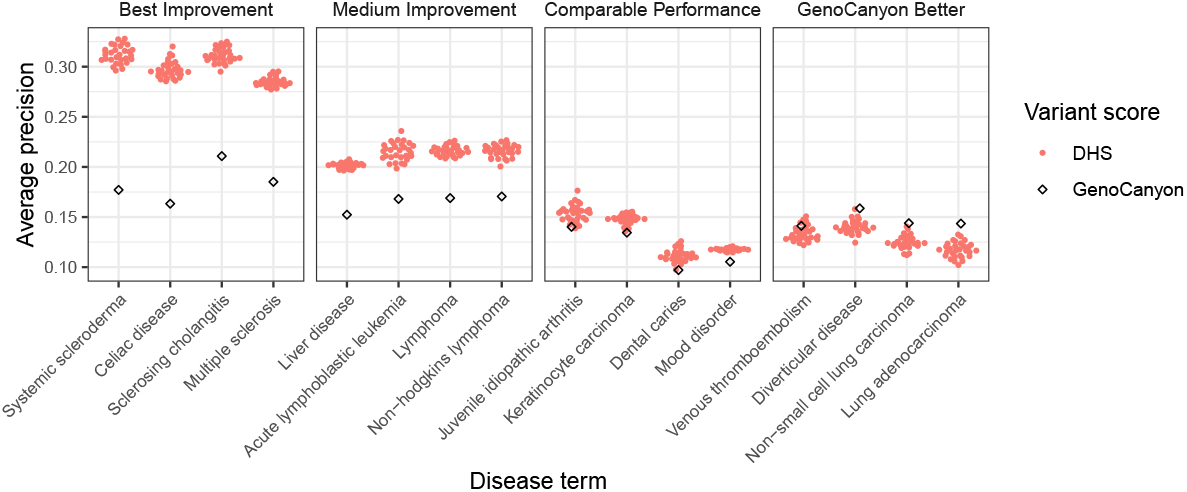
DHS disease-specific tissue weights improve variant prioritization compared with organism-level scores. For four strata (best improvement, middle improvement, comparable performance, and worse performance) we selected four disease terms and compared performance results. GenoCanyon (best organism-level score) performance is denoted in black, DHS tissue-weighted in red. Different performances of DHS tissue-weighted represent variation different data splits during nested cross validation (see **Methods**).

To make DHS tissue-weighted scores available, we generated pre-computed scores for 111 diseases at every base across the genome (for chromosomes 1-22, available at https://doi.org/10.7910/DVN/AUAJ7K). Scores were calculated at 25 bp resolution, the same as DHS scores.

### 2.4 DNase I hypersensitivity (DHS) scoring performs well compared with DIVAN

Here we compare the performance of tissue-weighted DHS scoring with DIVAN [19], a disease-specific variant score for 45 diseases. DIVAN is based on a more complicated feature-selection and ensemble-learning framework, and it uses a variety of other functional genomics features, in addition to DNase I hypersensitivity. To compare our method with DIVAN, we mapped EFO disease terms to MeSH terms (as used by DIVAN) and use MeSH terms for this section (See **Suppl. Data** SD12). Because DIVAN uses as supervised learning approach, and because the published model was trained using GWAS SNVs, it was necessary to create specific train and test datasets to ensure a meaningful comparison between tissue-weighted DHS and DIVAN.

Therefore, to assess performance of both DIVAN and DHS, we created a test set of disease-associated variants (and their matched controls) that were published later than 2016 (DIVAN’s publication date). That is, these variants are unlikely to have been a part of DIVAN’s training data. We also created a training set for DHS tissue-weightd containing only SNVs published prior to 2016. This resulted in training data that (a) is distinct from the test set and (b) draws on similar information that was available for DIVAN’s training. Further on, we only selected disease terms for this training/test data combination where at least 20 term-associated SNVs were present in the training data, and where at least 50 SNVs were present in the test data. This approach yielded 29 disease terms for this analysis. We then re-trained tissue-weighted DHS on this training data and compared with DIVAN on the test data. In addition, we added the organism-level GenoCanyon score as a reference.

To assess performance, we performed all pairwise comparisons for each disease term, and evaluated performance based on average precision. **Tab**. 4 summarizes observations, where we find that DHS performs significantly better than GenoCanyon and DIVAN in a majority of comparisons; however, there is a substantial number of comparisons (22 out of 58) where either GenoCanyon or DIVAN outperform DHS. **Fig**. 6 further illustrates these comparisons. In panel **A** we show performance across disease terms, grouped by the best-performing method. We see that tissue-weighted DHS outperforms DIVAN and GenoCanyon substantially on Multiple Sclerosis (MeSH:D009103), Psoriasis (MeSH:D011565) and Inflammatory Bowel Disease (MeSH:D015212); DIVAN outperforms GenoCanyon and DHS on Arthritis, rheumatoid (MeSH:D001172) and Heart failure (MeSH:D006333); GenoCanyon outperforms DHS and DIVAN on Stroke (MeSH:D020521) and Alzheimer disease (MeSH:D000544). In panels **B-D** we directly summarize comparison results; we observe that the DHS tissue-weighted score often has an advantage in terms where prioritization efforts are overall more successful (upper right quadrants). Finding overall good performance for our approach, we next more closely examined the disease-specific tissue aggregation weights we derive with our approach.

**Figure 6.**
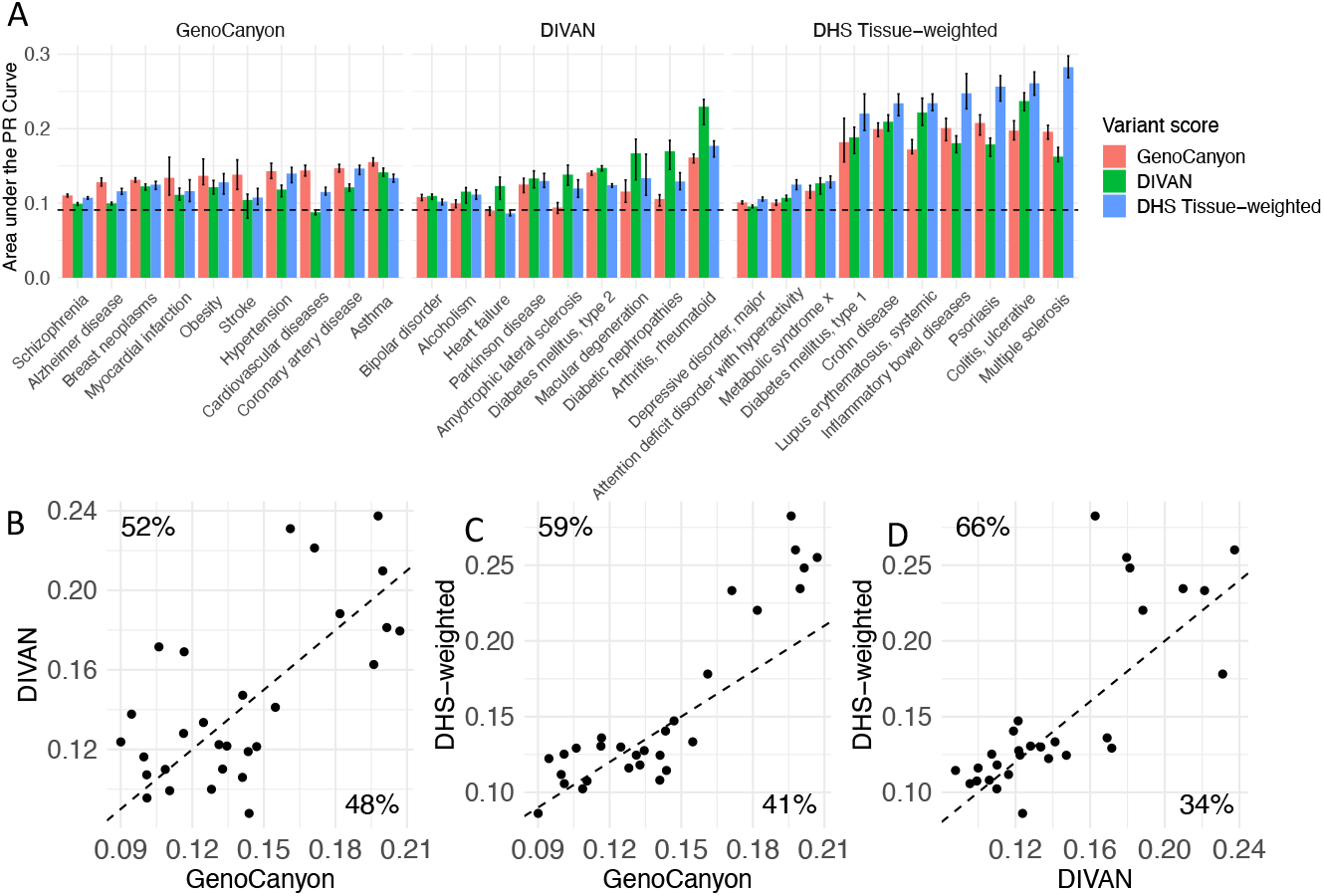
DHS tissue-weighted scoring outperforms DIVAN. Performance of DIVAN, Geno-Canyon, and DHS tissue-weighted across a test set, with disease terms grouped by the best-performing method. Vertical striped indicates the minimum and maximum performance of 30 bootstrap samples (A). Performance scatter plots of GenoCanyon vs. DIVAN performance (B); GenoCanyon vs. DHS-weighted (C); DIVAN vs. DHS-weighted performance (D). Average precision was used for these plots; dashed lines denote equal performance. Percentages denote the fraction of points above and below the diagonal, respectively.

**Table 4:** DHS tissue-weighted disease-specific scoring outperforms DIVAN. Across 29 disease terms, this table summarizes all pairwise comparison for DHS tissue-weighted, GenoCanyon and DIVAN using a specifically created test dataset. Wins, losses, and ties refer to significantly better (or worse, or tied) performance. Average precision was used as the performance metric, and the Wilcoxon singed-ranks test to determine wins and losses (p-values less than 0.05 were ties). Winning percent = #Wins/(#Wins+#Losses)

| Score | Wins | Losses | Ties | Winning percent |
| --- | --- | --- | --- | --- |
| DHS | 34 | 22 | 2 | 61 |
| GenoCanyon | 26 | 31 | 1 | 46 |
| DIVAN | 25 | 32 | 1 | 44 |

### 2.5 Disease-specific tissue weights reflect biomedical relevance

In addition to prioritizing SNPs, we can interpret the disease-specific tissue weights that our model learns in the context of disease mechanisms. Specifically, large tissue weights implicate tissues with a prominent role in associating SNVs with a disease in our model; therefore, one may hypothesize that such tissues or cell-types have a function in the etiology of that disease. To investigate this hypothesis, we analyzed tissue weights of the top-performing models we derived, where each model represents a different disease.

Results are summarized in **Tab**. 5; they include the two top-performing models, Systemic scleroderma (rank 1) and Sclerosing cholangitis (rank 2). In order to report a diverse range of diseases, we next excluded any diseases that are descendants of immune system disease (EFO:0000540) or lymphoma (EFO:0000574). From the remaining diseases, we identify the next three highest-ranked models: Colorectal adenoma (rank 15), Atrial fibrillation (rank 20), and Cutaneous melanoma (rank 21). For each diseases, we list the five tissues with the largest tissue-weights, and their tissue group.

**Table 5:** Top-ranked tissues for five diseases. For five diseases when show the top-five tissues with the largest tissue weights in the corresponding model we derive. The first column is the tissue rank, the second the tissue’s roadmap ID, the third the tissue name, the fourth the tissue group, and the fifth listst the adjusted p-value in an enrichment analysis performed by epimap [15].

| Rank | ID | Tissue name | Group |
| --- | --- | --- | --- |
| Systemic sclerosis |  |  |  |
| 1 | E116 | GM12878 Lymphoblastoid Cells | blood |
| 2 | E032 | Primary B cells from peripheral blood | blood |
| 3 | E041 | Primary T helper cells PMA-I stimulated | blood |
| 4 | E123 | K562 Leukemia Cells | blood |
| 5 | E030 | Primary neutrophils from peripheral blood | blood |
| Sclerosing cholangitis |  |  |  |
| 1 | E116 | GM12878 Lymphoblastoid Cells | blood |
| 2 | E061 | Foreskin Melanocyte Primary Cells skin03 | skin |
| 3 | E102 | Rectal Mucosa Donor 31 | gi_rectum |
| 4 | E041 | Primary T helper cells PMA-I stimulated | blood |
| 5 | E029 | Primary monocytes from peripheral blood | blood |
| Colorectal adenoma |  |  |  |
| 1 | E102 | Rectal Mucosa Donor 31 | gi_rectum |
| 2 | E110 | Stomach Mucosa | gi_stomach |
| 3 | E057 | Foreskin Keratinocyte Primary Cells skin02 | skin |
| 4 | E101 | Rectal Mucosa Donor 29 | gi_rectum |
| 5 | E028 | Breast variant Human Mammary Epithelial Cells (vHMEC) | breast |
| Atrial fibrillation |  |  |  |
| 1 | E083 | Fetal Heart | heart |
| 2 | E108 | Skeletal Muscle Female | muscle |
| 3 | E107 | Skeletal Muscle Male | muscle |
| 4 | E088 | Fetal Lung | lung |
| 5 | E120 | HSMM Skeletal Muscle Myoblasts Cells | muscle |
| Cutaneous melanoma |  |  |  |
| 1 | E061 | Foreskin Melanocyte Primary Cells skin03 | skin |
| 2 | E059 | Foreskin Melanocyte Primary Cells skin01 | skin |
| 3 | E117 | HeLa-S3 Cervical Carcinoma Cell Line | cervix |
| 4 | E041 | Primary T helper cells PMA-I stimulated | blood |
| 5 | E122 | HUVEC Umbilical Vein Endothelial Primary Cells | vascular |

The tissues we associate with disease, overall, appear reasonable and generally are in-line with existing knowledge about disease mechanisms. Systemic scleradoma is an autoimmune disorder that can affect skin and internal organs [24]. We find that GM12878 lymphoblastoid cells (a type of B cell) are among highest-weighted tissues, as were other types of B cells (primary B cell and B cell lymphoma, respectively). This in-line with previous studies that have shown that B cells play a role in system scleroderma [25, 26]. Sclerosing cholangitis is an inflammatory condition that leads to scarring and narrowing of the bile ducts [27]. We highlight various inflammation-related types of blood cells, such as T cells and monocytes, which were previously suggested to play a role in the disease [28]. Colorectal adenoma is a benign tumor that develops in the lining of the colon or rectum. Our model identified rectal mucosa and stomoch mucosa as the most-highly weighted tissues, and the function of rectal mucosa in colorectal cancer has been previously studied [29]. While the direct relationship between other gastrointestinal tissues and the development of colorectal adenoma has not been established, the association between gastrointestinal microbiome and colorectal adenomas has been discovered [30]. Regarding atrial fibrillation, our approach highlights fetal heart and lung tissues. In addition, we identified skeletal muscle cells. In the case of cutaneous melanoma, a type of skin cancer, our approach emphasizes foreskin melanocyte cells and a specific type of T cell. Apart from these, we highlight cervical carcinoma cell lines and endothelial primary cells.

Overall, we conclude that the tissue weights we derive carry biomedically meaningful information and are able to highlight tissue contexts that may play a role in disease etiology. To further explore this finding, we used a resource of the epimap consortium [15], where disease-tissue associations are reported that derived differently from the one we obtained in two key ways: First, epimap uses their enhancer definitions based on a much larger set of genome annotations. Second, epimap’s enrichment test contrasts disease-associated SNP enrichment in a specific tissue’s enhancer set compared to all enhancers, whereas our method effectively compares open chromatin harboring disease-associated SNPs vs control SNPs tissue-by-tissue. Nevertheless, results are summarized in **Suppl. Data** ST7, and we find that out of the 25 tissues we associate with disease terms 14 have an estimated false discovery rate of less than 4% in the epimap analysis as well. Notably, a ground truth for these association is generally unknown; but we interpret the overlap in associations as encouraging, while complementary associations are expected, given the differences in methodology. Based on this overall finding of meaningful disease-tissue associations, we next further explored the use of tissue-weights in disease charactrization.

### 2.6 Disease-term similarity based on DHS tissue-weighted modeling reveals meaningful groups

Disease-specific tissue weights for aggregating DHS scores, which are learned by our approach, can highlight tissues and cell-types with a role in the disease (see previous section). Therefore, we derived and explored a measure for disease similarity based on these weights.

#### 2.6.1 Disease similarities based on disease-specific tissue weights for non-coding variant prioritization

In our DHS tissue-weighted approach, for each disease term DNA accessibility across the same set of tissue and cell-type contexts is used to predict whether a certain SNV is disease-associated, or not. This results in disease-specific tissue aggregation weights (that is, coefficients in our logistic regression model)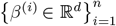, where *i* is indexing disease terms, *n* is the number of disease terms studied, and *d* denotes the number of tissues/cell-types with DHS scores. For our similarity measure between two diseases, say *i* and *j*, we then use a version of the Pearson correlation between *β*^(*i*)^ and *β*^(*j*)^that takes uncertainty in the estimated aggregation weights into account (see **Methods**). That is, if an overlapping set of tissues/cell-types drive the prioritization of SNVs for two diseases, similarity is high; if different tissues are used, similarity is low.

Using this approach we calculated disease similarities for the 111 disease terms we study. Resulting similarities are visualized in (**Fig**. 7), where we show a similarity-based two-dimensional UMAP projection of disease terms. We observe that disease terms segregate into separate groups, with a coarse grouping between immune related diseases (lower left inlay, black) and others (lower left inlay, gray). A higher-resolution group structure was obtained by sub-clustering, where we grouped disease terms into seven groups (main panel, **Fig**. 7). Clusters names are based on EFO disease terms that include a large amount of cluster members as child-terms (see **Methods** and **Suppl. Fig**. S10-S16); **Tab**. 6 lists disease terms per cluster. In addition to the clear separation of immune-related diseases from others, we also find a very homogeneous group consisting of mental and behavioural disorders, containing terms like schizophrenia (EFO:0000692) and anxiety disorder (EFO:0006788), and a group of skin cancers. The remaining three groups are more heterogeneous, but with two of them containing several terms related to cardiovascular disease (EFO:0000319) and digestive system disorders (EFO:1000218), respectively. By design similar tissues in each group drive SNP-disease associations, and we next examined which tissues play a role in each of the clusters.

**Figure 7.**
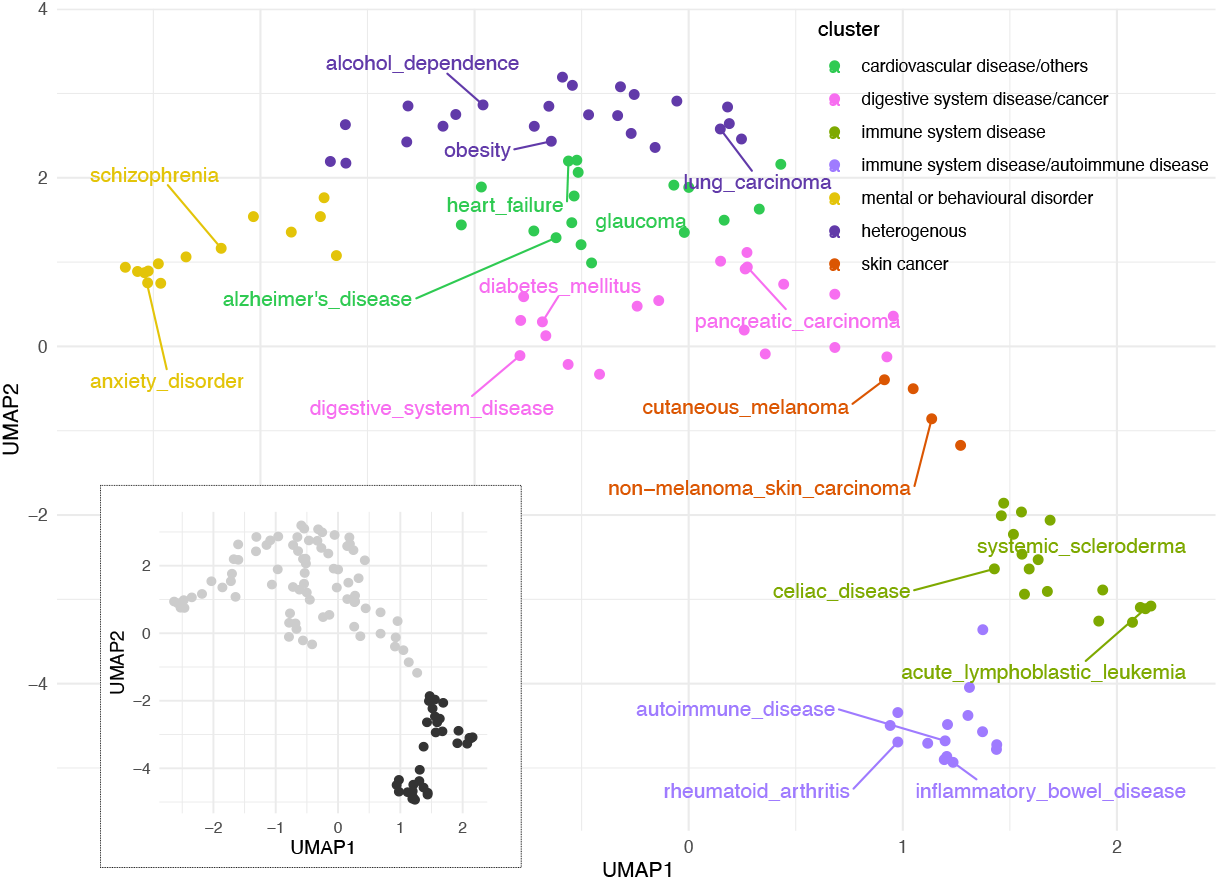
Similarity-based two-dimensional projection visualizes 111 diseases. Two dominant disease groups emerge in this visualization (immune system related disease terms (black) and others (gray), in the inlay). Hierarchical clustering was used to group diseases into seven clusters, with colors indicating broad disease types (see **Tab**. 6 for details).

**Table 6:**
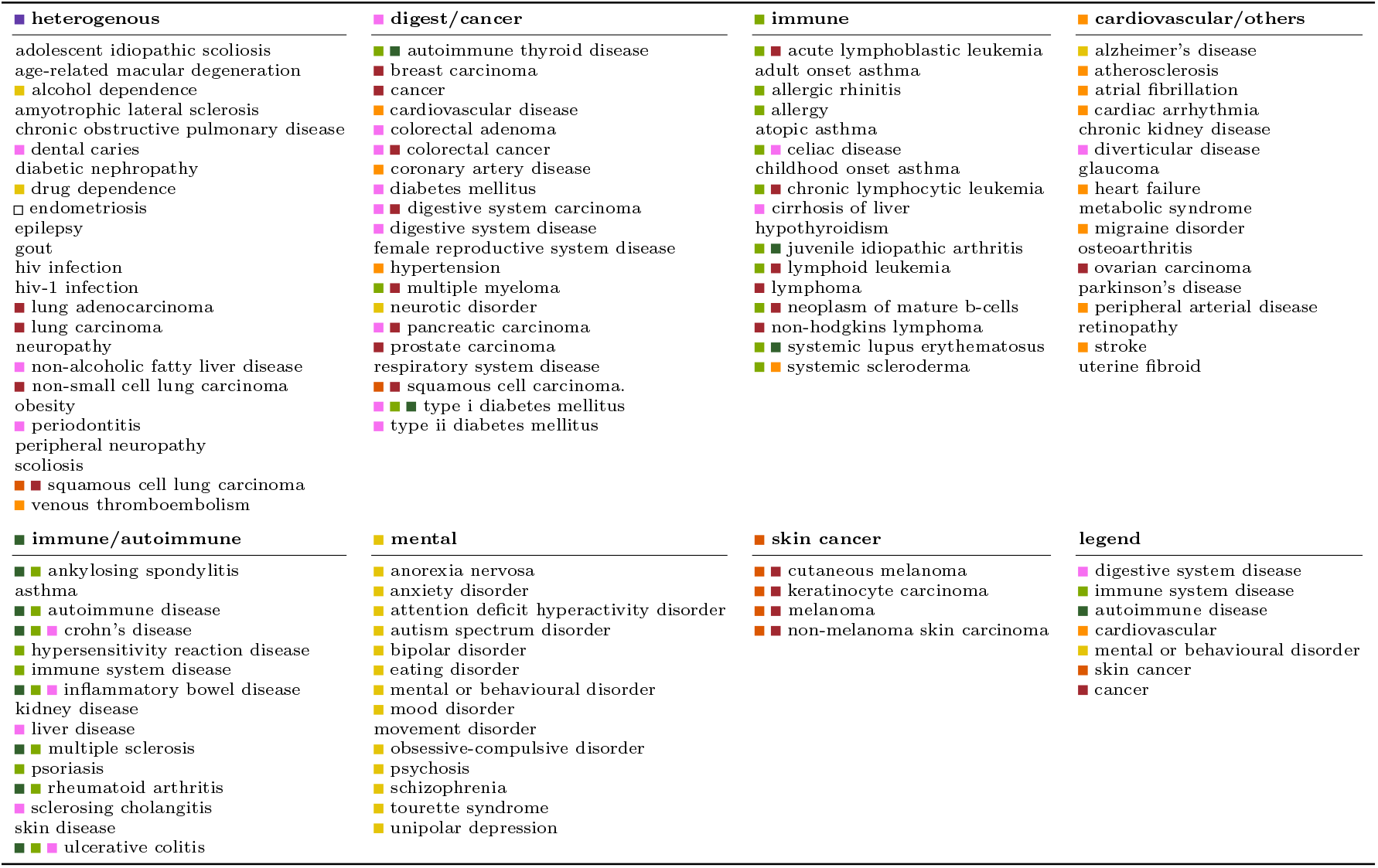
Disease groups based on DHS tissue-weights. For each disease group disease terms are shown. The colored squares denote the disease groups in the EFO ontology.

**Table 7:** Example disease pairs of genetic correlation and model similarities. This table shows the genetic correlation and model similarity for some disease pairs as we selected. *s*_*g*_: genetic correlation; *s*_*m*_: model similarity. For quadrant B, C, D we pick 8 disease pairs, where *s*_*g*_ + *s*_*m*_, *s*_*g*_ − *s*_*m*_ and *s*_*m*_ − *s*_*g*_ are the highest, respectively.

| Disease 1 | Disease 2 | $s_g$ | $s_m$ | Quadrant |
| --- | --- | --- | --- | --- |
| Inflammatory bowel disease | Ulcerative colitis | 1.00 | 0.88 | B |
| Diabetes mellitus | Type ii diabetes mellitus | 0.91 | 0.91 | B |
| Crohn's disease | Inflammatory bowel disease | 0.72 | 0.91 | B |
| Sclerosing cholangitis | Ulcerative colitis | 0.63 | 0.82 | B |
| Crohn's disease | Ulcerative colitis | 0.53 | 0.84 | B |
| Ankylosing spondylitis | Sclerosing cholangitis | 0.35 | 0.90 | B |
| Inflammatory bowel disease | Sclerosing cholangitis | 0.44 | 0.76 | B |
| Bipolar disorder | Schizophrenia | 0.71 | 0.42 | B |
| Rheumatoid arthritis | Systemic lupus erythematosus | -0.47 | 0.51 | C |
| Celiac disease | Systemic lupus erythematosus | -0.16 | 0.58 | C |
| Sclerosing cholangitis | Systemic lupus erythematosus | -0.24 | 0.49 | C |
| Crohn's disease | Sclerosing cholangitis | 0.17 | 0.83 | C |
| Rheumatoid arthritis | Sclerosing cholangitis | 0.07 | 0.69 | C |
| Crohn's disease | Rheumatoid arthritis | 0.06 | 0.66 | C |
| Systemic lupus erythematosus | Ulcerative colitis | -0.16 | 0.43 | C |
| Crohn's disease | Systemic lupus erythematosus | -0.10 | 0.49 | C |
| Type i diabetes mellitus | Type ii diabetes mellitus | 0.85 | 0.10 | D |
| Diabetes mellitus | Type i diabetes mellitus | 0.91 | 0.20 | D |
| Celiac disease | Obsessive-compulsive disorder | 0.36 | -0.26 | D |
| Diabetes mellitus | Obesity | 0.54 | 0.01 | D |
| Obesity | Osteoarthritis | 0.49 | 0.02 | D |
| Attention deficit hyperactivity disorder | Obesity | 0.44 | 0.03 | D |
| Attention deficit hyperactivity disorder | Osteoarthritis | 0.40 | 0.00 | D |
| Obesity | Type i diabetes mellitus | 0.40 | 0.00 | D |

In order to find group-specific tissues, we examined for each cluster the top five tissues that (a) contribute most to disease association and (b) are cluster specific (see **Methods**). Results are summarized in **Fig**. 8; we note that both disease groups related to the immune system highlight blood tissues (such as E043: Primary T helper cells from peripheral blood and E116: GM12878 Lymphoblastoid Cells, see **Suppl. Data** SD23 for all names of standard epigenomes), with the group containing inflammatory bowel disease, Crohn’s disease, and ulcerative colitis also containing rectum tissues (such as E101: Rectal Mucosa Donor 29). Brain tissues contribute to disease associations for mental and behavioral disorders, skin tissues to skin cancer, and gastro-intestinal / stomach tissue to the cluster with digestive system diseases. We also note that a clear association of specific tissues with a disease group correlates with better classification performance of our model for SNP-disease association (**Fig**. 8; for example, see the immune and immune/autoimmune clusters). We note, though, that not for all clusters the corresponding tissue associations are equally compelling, as illustrated in the same figure. While the clusters we derive resemble broader disease groups, for each disease a specific combination of tissues is used to derive whether a variant might be associated, and some tissues contribute to several clusters. For instance, one blood cell type (E116, GM12878 Lymphoblastoid Cells) contributes to both immune clusters, but also to diseases in the digestive/cancer, heterogeneous and skin cancer clusters. Another blood cell type (E043, Primary T helper cells from peripheral blood) displays a similar pattern. **Suppl. Fig**. S9 shows the same heatmap as **Fig**. 8, but for all tissues.

**Figure 8.**
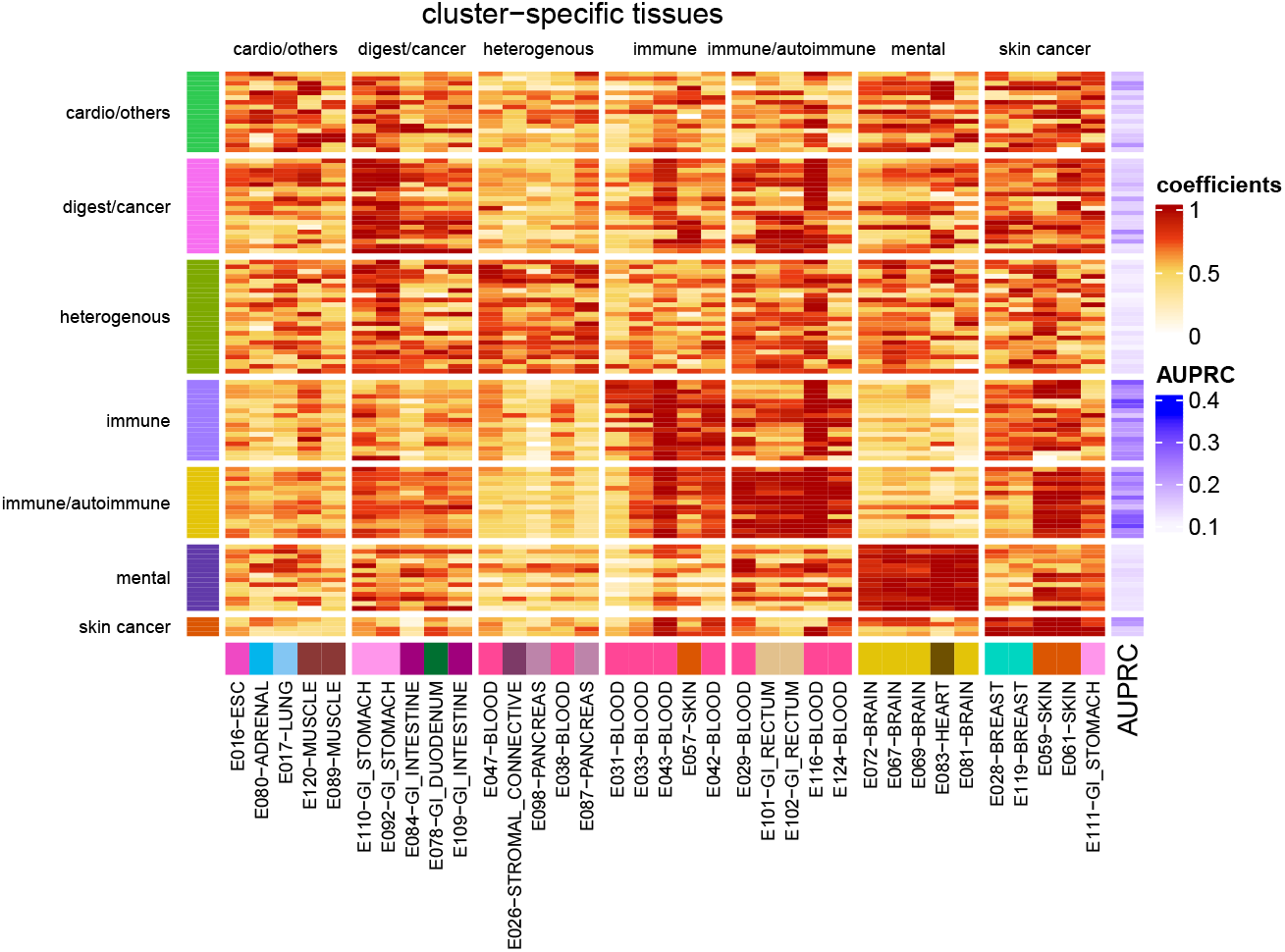
Heatmap of top-five tissue-weights for 111 diseases. Regularized model coefficients (i.e., tissue weights) of five disease-cluster-specific tissues (columns) are shown for 111 diseases (rows). Coefficients are scaled by disease, and rows are grouped into sets of cluster-specific tissues (see **Methods** section). Bottom annotation shows tissue names of cluster-specific tissues (names are shown in the format of ‘Tissue name’ -’Tissue group’; annotation on the left side shows disease cluster, and annotating on the right side shows model performance in terms of AUPRC).

Overall, these results suggest that our modeling approach successfully identifies tissues with a role in disease etiology. Finally, we explore how our disease similarities relate to genetic similarities as measured by genetic correlation between diseases.

#### 2.6.2 Model-based similarities are complementary to genetic correlation

Here we compare the disease-disease similarities we derived (*s*_*m*_) with genetic correlations from the GWAS Atlas (*s*_*g*_), where genetic correlation measures shared genetic causes between two traits [31]. For 6,105 possible disease pairs of the 111 diseases terms we study, estimates of genetic correlation for 595 pairs were available from the GWAS Atlas (see **Methods**). Overall, for these 595 disease pairs we observe only weak (but statistically significant) correlation between model similarities and genetic correlations (*r* = 0.32, *p value* = 2.4*E* − 15), where the scatter plot is shown in **Fig**. 9 panel A.

We also see that most disease pairs are not annotated with substantial genetic correlations, or with high model-based similarities (90% of disease pairs have *s*_*m*_ *<* 0.25, and *s*_*g*_ *<* 0.2). Therefore, we explored three different regimes: Disease pairs where both similarity measures are high (*s*_*m*_ ≥ 0.25 and *s*_*g*_ ≥ 0.20), pairs with high genetic correlations and low model similarity (*s*_*m*_ *<* 0.25 and *s*_*g*_ ≥ 0.20) vice versa (quadrants indicated in **Fig. 9A**, named quadrants B, C and D). The top eight most extreme examples from each regime are summarized in **Tab**. 7. In the following we discuss some examples in more detail. Specifically we explore two immune system diseases for quadrant B; two mental or behavioral disorders for quadrant C; and one immune system disease and one mental or behavioral disorder for quadrant D. We note that the pairs we examine have no annotated parent-child relationships in the EFO.

- Ulcerative colitis (UC, EFO:0000729) and Crohn’s disease (CD, EFO:0000384) have both high genetic correlation (*s*_*g*_ = 0.53) and model similarity (*s*_*m*_ = 0.84), see **Fig. 9A**. This suggests that they share genetic causes, and that the same tissues are informative for SNP-disease association. While shared genetic causes for UC and CD have been pointed out (e.g., [32]), our model for SNP-disease association allows us to explore relevant tissue contexts. In **Fig. 9B** we show a scatter plot of tissue weights for both diseases, where color indicates the importance of each tissue to model similarity (see **Methods**). We observe that open chromatin in blood (E116, GM12878 Lymphoblastoid Cells; E124, Monocytes-CD14+ RO01746 Primary Cells; E041, Primary T helper cells PMA-I stimulated) and rectum (E102, Rectal Mucosa Donor 31) is positively associated with SNP-disease association in both diseases; this is consistent with a previous study where blood cell types are found to be relevant in many autoimmune diseases, including UC and CD [33]. In addition, symptoms or complications in rectum is also observed in UC and CD [34]. Interestingly, open chromatin in GI-intestine (E085, fetal intestine small) is negatively associated with SNP-disease association, along with ohter intestine tissues (E084, fetal intestine large and E109, small intestine, with the 61th and 86th smallest tissue weight, respectively, amongst 127 contexts). This indicates fetal intestine or small intestine might be less involved in UC and CD etiology, compared to their juvenile and adult counterparts.
- Autism spectrum disorder (ASD, EFO:0003756) and anorexia nervosa(AN, EFO:0004215)] is an example where we observe a low genetic correlation (*s*_*g*_ = −0.05) and a moderate high model similarity (s_m_ = 0.34); a scatter plot of their tissue weights is shown in **Fig**. 9C. Note that we did not choose one of the highlighted pairs in **Tab**. 7 for this quadrant, because we already discussed a immunesystem realted disease pair. We observe that both disease models give heart and brain tissue (E083, fetal heart and E081, fetal brain male) high tissue weights. This is consistent with the observation of brain abnormalities in ASD and AN [35, 36]. While the presence of fetal heart is less intuitive, we note that children with abnormal heart development are more likely to develop ASD, suggesting a connection between the disease and the fetal heart [37]. We also note that while genetic correlation between ASD and AN is low, a link between the two diseases on the phenotypic level is being suggested [38, 39]; the tissue context we identified could provide information about shared molecular aspects of disease etiology as well.
- For obsessive compulsive disorder (EFO:0004242) and celiac disease (EFO:0001060) we observe low model similarities (*s*_*m*_ = −0.26) and moderately high genetic correlation(*s*_*g*_ = 0.36); **Fig**. 9 **D** shows the scatter plot of tissue weights. Several studies have shown that nervous system disease and immune related diseases have shared genetic background [40, 41]. However, in contrast to the other two examples, there is little relation between tissue weights in these two diseases. Blood cell types are highlighted in celiac disease, while brain and fetal heart tissues are highlighted in obsessive compulsive disorder. For celiac disease, the top six tissue contexts are blood cells, including different types of T cells (E041, Primary T helper cells PMA-I stimulated; E043, Primary T helper cells from peripheral blood and E034, Primary T cells from peripheral blood) and lymphoblasts (E116, GM12878 Lymphoblastoid Cells), which is consistent with findings that alterations in T cells and lymphoblasts can lead to celiac disease [42, 43].

**Figure 9.**
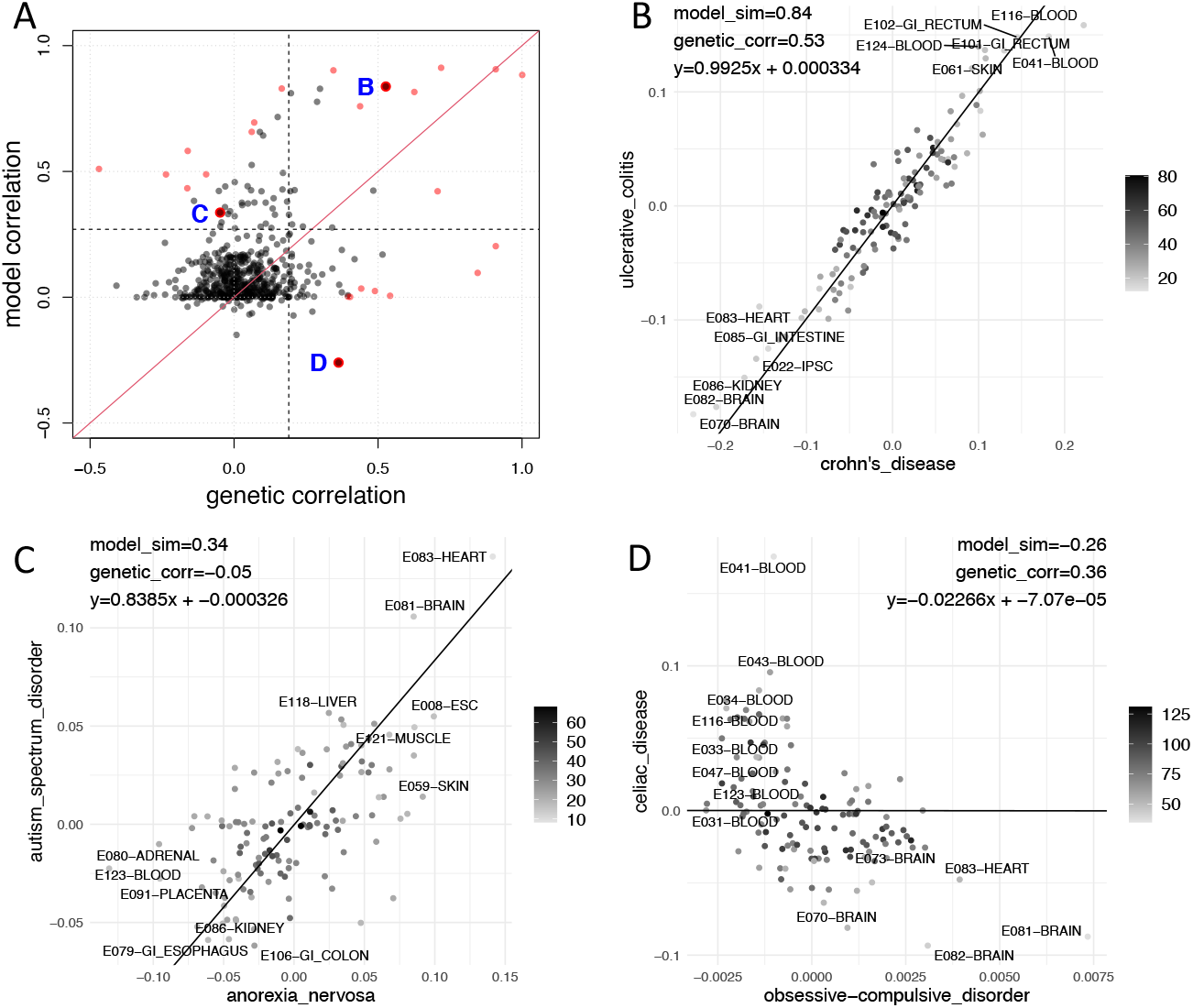
Genetic correlation and model similarity. *(A) Genetic correlation vs. model similarity for 595 disease pairs*. Each point is a disease pair, where the *x*-axis denotes the genetic correlation and *y*-axis is the disease model similarity. For three quadrants we highlight disease pairs, denoted by B, C, and D). (B-D) Scatter plot of tissue coefficients in three example disease pairs, where (B) shows Crohn’s disease vs inflammatory bowel disease; (C) shows anorexia nervosa vs autism spectrum disorder and (D) shows celiac disease vs obsessive compulsive disorder. Lines denote a weighted linear regression line underlying our disease similrities. Color codes for the weight for each tissue when conducting weighted regression analysis.

Overall, these examples illustrate that the disease similarities we derive are complementary to genetic correlation. In addition, tissue contexts highlighted by our tissue-weights allow for biomedical interpreations of observed similarities (i.e., which are the relevant tissue contexts) and can be used to generate molecular hypotheses about disease etiology.

In summary, our results show that disease-specific variant prioritization performs well for non-coding GWAS variants, compared with organism-level approaches. We also demonstrate that disease-specific tissue-weights are biomedically meaningful and can be used to generate hypotheses about disease mechanism. Therefore, we believe this type of variant characterization is a useful tool for researchers studying the molecular and genetic causes of disease.

## 3 Discussion

Most variant scores prioritize non-coding variants either at the level of the whole organism (e.g, CADD [8], GenoCanyon [44]), or they provide tissue-specific scores (e.g, GenoSkyline [11], Fitcons2 [12]). Here we present a straightforward strategy to combine tissue-specific variant scores in a disease-specific manner. We show that for common genetic variants in the GWAS catalog [1] our approach leads to better performance than organism-level or tissue-specific scores (see **Fig**. 5). Pre-computed disease-specific prioritization scores are available at https://doi.org/10.7910/DVN/AUAJ7K.

Comparing different variant prioritization methods we note that we use area under the precision-recall curve as an evaluation metric, and that the performance of all methods is modest. We believe that is because our analysis ***(a)*** focuses explicitly on non-coding variants, ***(b)*** stratifies SNVs by disease-phenotype, and ***(c)*** utilizes unbiased matching of control-SNVs (SNPsnap-matching, see Section 4.1.2). Each of these points affects the SNV sets we use for our analysis, and therefore the performance metrics we report. For transparency we provide all disease-associated variants we use (with matched negatives) in our supplemental data. As a more general point we also note that associations reported in the GWAS catalog contain causal as well as non-causal SNPs, which will also contribute to sub-optimal performance measures of all the variant scores we assess.

We included a comparison with the DIVAN method in our evaluation, which also includes comparing GenoCanyon with DIVAN. Part of this comparison is analogous to results reported in Chen et al. [19]; however, the performances we observed do not agree perfectly, as detailed in **Suppl. Data** SD15. Broadly, looking at overlapping/matching disease terms, our results appear more favorable for GenoCanyon. These differences are likely due to different test sets used in the two evaluations (i.e., the GWAS catalog (this study) vs. GRASP).

We also note that there is other research associating variants with disease terms in a similar setting, notably PINES [20] and LSMM [45]. We did not compare directly with PINES, because no pre-computed scores are available; also, we note that while performance reported in this publication in terms of AUROC is higher than our results, a less stringent un-matched test set of random/control variants was used in these analyses. For LSMM we note that we leverage variants associated with EFO disease terms across studies, while LSMM uses summary statistics on a per-study basis. Using aggregate data from different studies allows our approach to consider parent-child relationships of the EFO ontology using variant aggregation (see Section 2.1).

We demonstrate that our approach can be used to calculate similarities between disease terms, see Section 2.6.1. Since this similarity measure is derived from non-coding SNVs associated with disease, one could expect it is largely congruent with genetic correlation between disease traits. However, that is not the case (see **Fig**. 9), most likely because we focus on a small subset of disease-associated SNVs reported in the GWAS catalog. For example, obsessive-compulsive disorder and celiac disease have a high genetic correlation (*s*_*g*_ = 0.36) but do not share noncoding SNPs in the GWAS catalog (and low model similarity *s*_*m*_ = −0.26); on the other hand, autism spectrum disorder and anorexia nervosa have a low genetic correlation (*s*_*g*_ = −0.05) but share a number of significant SNPs in the GWAS catalog (and relative high model similarity *s*_*m*_ = 0.34). In addition, interpretation of model similarity between disease terms is different from genetic correlation; high model similarity implies that disease-associated SNVs reside in DNA-accessible regions in an overlapping set of tissues, but the identity of individual SNVs (and whether they overlap) is inconsequential. For example, asthma and rheumatoid arthritis have only 15 shared SNPs (out of 732 and 1283 SNPs in rheumatoid arthritis and asthma, respectively), but exhibit high model similarity (*s*_*m*_ = 0.53). This shows that model similarity between two diseases can involve similar tissues even if they do not share a genetic background. Further on, we note that estimates of genetic correlation also may depend on the study used. For example, systemic lupus erythematosus (SLE) has a negative genetic correlation (*s*_*g*_ = −0.47) with rheumatoid arthritis (RA) (and other inflammatory diseases) when using the SLE summary statistics from Julia et al. [46] (as retrieved from the GWAS Atlas [31]), whereas another study (Lu et al., [47]) found SLE to have a positive genetic correlation (*s*_*g*_ = 0.41) with RA when using the SLE summary statistics from Bentham et al. [48].

We note that in our analyses we used the EFO ontology to aggregate variants annotated in the NIH/EBI GWAS catalog. That is, for each disease term directly-annotated variants were used, and, in addition, variants annotated to descendant terms in the ontology were also included. This approach allowed us to compile a more exhaustive set of variants per term. However, some amount of caution should be exercised when using disease models with more general terms, such as “cardiovascular disease” for example, as they may encompass heterogeneous diseases.

Our approach is expected to improve as more variants are associated with disease, and as disease-associations get more refined. In addition, increasing amounts of epigenomics data, such as epimap [15] and ENCODE5 [6], could be incorporated and they have the potential to improve the disease associations we learn.

In summary, we have provided a straightforward method to leverage tissue-specific variant scores for disease-specific variant prioritization. We show that this approach performs well compared with current methods, and we show that the resulting association models are interpretable and lead to useful characterization of disease terms. Overall, our contributions are useful for the following two reasons: Conceptually, because they highlight the value of disease-specific variant prioritization. In addition, we provide pre-computed association scores for 111 disease terms that researchers can use in practice to interpret their variant data.

## 4 Methods

### 4.1 Data sources and processing

#### 4.1.1 Disease-associated variants

Disease-associated non-coding single nucleotide variants were retrieved from the NHGRI-EBI Catalog of human genome-wide association studies database (GWAS catalog, version `2020-12-02`, downloaded from https://www.ebi.ac.uk/gwas/docs/file-downloads). These data contained 122,396 unique non-coding SNPs spanning 2,782 phenotypes, where non-coding was defined as variants not overlapping protein-coding sequence (GENCODEv36); we also excluded variants annotated as protein coding sequence variants (e.g. missense variants, frameshift variants) as a SNP’s “functional class” in the GWAS Catalog. Further, variants in the GWAS Catalog are annotated with phenotypes using the Experimental Factor Ontology (EFO, https://www.ebi.ac.uk/efo) [49]. We focused on variants with phenotype terms annotated in disease domain of the EFO (i.e., all terms/traits/phenotypes we consider are descendants of the term “disease” (EFO:0000408, EFO version `3.24.0`, accessed `2020-11-17`). Further on, SNPs in the HLA region, and SNPs with minor allele frequency (MAF) less than 1% in the European population as reported by the International Genome Sample Resource were excluded (as they cannot be matched to control SNPs with the SNPsnap approach, see below). Out of 31103 SNVs, a total of 5225 SNVs were removed. Finally, in our analyses we restricted ourselves to phenotypes with at least 100 annotated non-coding SNPs. **Suppl. Data** SD1 and SD2 contain 111 phenotypes and 77,028 phenotype-associated SNPs we used in this study. We also grouped SNPs in LD blocks (SNPsnap, *r*^2^ ≥ 0.5) and identify SNPs with the minimum p-value per block(“representative SNP”); we provide this information, which we use in some of the analyses described below, in **Suppl. Data** SD2.

#### 4.1.2 Control variants

For each disease-associated SNP we generated matched control non-coding variants MAF ≥1%) using four different strategies, where the non-coding is again defined discussed above (**Section** 4.1.1). The four strategies are:

##### Random

For each disease-associated SNP, we selected ten SNPs from common variants in 1000G EUR at random (i.e., equal probability for all SNPs) as controls.

##### TSS-matching

We processed common non-coding SNVs and selected a subset of these variants as controls, where the distribution of distances to the nearest protein-coding gene’s transcription start site (TSS) are matched between control set and disease-associated SNPs (similar to GWAVA, [23]). Specifically, we sorted all common non-coding SNPs by the distance to the nearest TSS and divided them into 50 bins, where each bin contains the same number of SNVs. Then, for each disease-associated SNP, we randomly selected ten control SNPs from the bin containing the disease-associated SNP’s distance to the nearest gene.

##### SNPsnap-matching

Using SNPsnap [22], we matched control SNPs to disease-associated variants in terms of minor allele frequency, gene density (distance cutoff `ld0.8`), distance to the nearest gene TSS, and number of SNPs in LD. Our parameters for maximum allowable deviation were: 5%, 50%, 20% and 50%, respectively. We randomly selected ten control SNPs per disease-associated SNP form SNPsnap’s results, and we ensured there are no duplicated control SNPs for different disease-associated SNPs. If there were less than 10 control SNPs returned by SNPsnap, we kept all of the control SNPs. If no control SNPs were matched, we removed the disease-associated SNVs (a total of 311 SNVs) from our analyses.

##### SNPsnap-TSS-matching

Essentially the same as in SNPsnap-matching, but controlling **only** for the distance to the nearest genes (maximum allowable deviation: 20%); for three other attributes “maximum allowable deviation” is set to 10,000%. We note that in both SNPsnap-matching and SNPsnap-TSS-matching, distance is measured by distance to the nearest gene, whereas for TSS-matching only protein-coding genes are considered.

In all four matching strategies we excluded variants annotated in the GWAS catalog as control SNPs. **Suppl. Data** SD3 contains the four sets of control variants.

#### 4.1.3 Additional data sources, variant scores

We used pre-computed SNP annotations from the following sources:

- CADD v.1.3: http://krishna.gs.washington.edu/download/CADD/v1.3/1000G_phase3.tsv.gz
- EigenPC v.1.1: https://xioniti01.u.hpc.mssm.edu/v1.1
- Fitcons2: http://compgen.cshl.edu/fitCons2/hg19
- GenoCanyon: http://genocanyon.med.yale.edu/GenoCanyon_Downloads.html
- GenoSkylinePlus: http://genocanyon.med.yale.edu/GenoSkylineFiles/GenoSkylinePlus/GenoSkylinePlus_bed.tar.gz
- GWAVA v.1.0: ftp://ftp.sanger.ac.uk/pub/resources/software/gwava/v1.0/VEP_plugin/gwava_scores.bed.gz
- LINSIGHT: http://compgen.cshl.edu/%7Eyihuang/tracks/LINSIGHT.bw
- DIVAN: https://sites.google.com/site/emorydivan
- DHS accessibility: We downloaded Avocado-imputed [50] DNase1 hypersensitive sites (DHS) signal for 127 ENCODE biological contexts (tissues / cell types) from https://noble.gs.washington.edu/proj/avocado/data/avocado_full/DNase/.

### 4.2 Tissue-weighted variant prioritization based on DNase1 hypersensitivity

#### 4.2.1 A penalized logistic regression model for context-weighted score averaging

For predicting SNP’s associations with a disease term, we consider SNPs as observations, and each SNP is described as a vector **x** ∈ ℝ^*d*^ of variant scores in *d* tissues/contexts; we arrange vectors 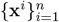 for *n* observations in a matrix *X* ∈ ℝ^*n×d*^, together with a vector *y* of *n* binary entries, indicating for each SNP association with a specific disease term (no=0/yes=1). In addition, we denote the average score (across contexts) for a SNP *i* by 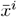, which is also a basline score because it aggregates across contexts.

We use a logistic regression model of the form

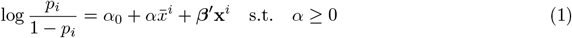

where *α*_0_ ∈ ℝ, *α* ∈ ℝ_+_ and ***β*** ∈ ℝ^*d*^ are regression coefficients, and *p*_*i*_ is the probability that SNP *i* is associated with a disease that is studied. We fit a regularized version of the negative log likelihood

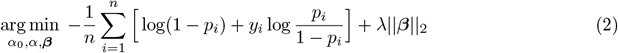

where the dependence on *α*, ***β*** of the first term is through Equation (1). For large regularization parameters *λ* this will yield small ***β*** → **0** and recover the baseline 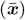 of unweighted averaging of context scores (scaled by a non-negative factor *α*). We implemented this approach using the R package glmnet (version 2.0-18, [51]) and determined the regularization parameter via 5-fold cross validation (cv.glmnet function) through maximizing the area under the (cross-validated) ROC curve. Class weights were employed to balance skewed class sizes.

#### 4.2.2 Disease similarities from context-weighted score averaging

Context-weighted score averaging, as described above, results in disease-specific coefficient vectors ({***β***^(*i*)^}, with *i* indexing disease terms), together with bootstrap estimates for the standard deviation of each coefficient (that can be arranged in corresponding vectors {***γ***^(*i*)^}). Specifically, we use 5-fold cross-validation repeated 10 times, yielding 50 coefficient vectors for each disease. We use their mean for our estimate of ***β***^(*i*)^, and their standard deviation as an estimate of ***γ***^(*i*)^.

For a pair of diseases (*d*_*i*_, *d*_*j*_) we then define a disease similarity through similarity of associated coefficient vectors ***β***^(*i*)^ and ***β***^(*j*)^, taking into account our estimates of coefficient variability. Specifically, we fit a weighted linear regression model (i.e., regressing ***β***^(*i*)^ on ***β***^(*j*)^), with regression weights taking into account coefficient variability as follows:

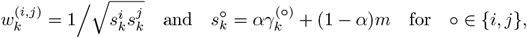

where we chose *m* to be the 25% quantile of all (esitmated) standard deviations observed, and *α* = 3/4. Therefore, 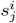 and 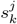 are shrunken versions of the standard deviations for the regression coefficients of disease *i* and disease *j* in tissue/context *k*, respectively. Finally, for disease pairs with a positive coefficient from the weighted linear regression we take the coefficient of determination (*R*^2^) as a similarity measure; for disease pairs with a negative coefficient, we take −*R*^2^. We note that for constant regression weights 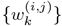 this is equal to the Pearson correlation between the coefficient vectors we obtain from context-weighted score averaging (i.e., cor(***β***^(*i*)^, ***β***^(*j*)^)).

### 4.3 Variant prioritization performance

#### 4.3.1 Tissue-weighted cross-validation performance

To measure the cross-validation performance of Tissue-weighted, we use repeated cross-validation [52] to reduce the variance (due to the random partitioning of data into 5 folds). Here, we repeated 5-fold cross-validation 30 times, and record the performance of each repeat. We later use the mean performance of the 30 repeats as the performance of that method and we also show the variance in figures such as **Fig**. 4.

#### 4.3.2 Comparing organism level scores

For each disease we have disease-associated and control SNVs, and corresponding pre-computed organism-level scores. With this setup we calculate performance metrics of interest (area under the receiver operator characteristic curve (AUROC) and average precision (AUPR)), and obtain disease-specific performance metrics for each scoring approach. To compare performance between organism-level scores on the same disease we use performance measures computed on 30 bootstrap samples (each bootstrap sample randomly contains 90% of disease and control variants) and then employ the Wilcoxon signed-ranks test to test to assess differences in performance. This yields p-values as reported in **Suppl. Data** SD4.

With respect to aggregating comparisons across diseases, we note that disease terms can (and do) share SNVs, so performance metrics in different terms are not necessarily independent. Also, disease terms can vary substantially in the number of annotated SNPs. We again use Wilcoxon singed-ranks test [53] on performance metrics (computed using all disease-associated-and control-SNVs for each disease term) to compare two organism-level variant scores aggregate across diseases. This approach yields p-values, as reported in **Suppl. Data** SD5.

#### 4.3.3 Comparing tissue-weighted scores

Tissue-weighted baseline scores (see above) are calculated in the same way as organism-level scores. For tissue-weighted scores with data-driven tissue-specific weighting (see above), we use cross-validated performance measure for each bootstrap sample and the same 30 bootstrap samples as when we compared between organism-level scores. And then we use the same Wilcoxon signed-ranks tests to measure the difference. For comparing scores aggregated across diseases we again proceed analogous to organism-level scores and use a Wilcoxon singed-ranks test on cross-validated disease-specific performance measures. Results are summarized in **Suppl. Data** SD8 and SD9.

#### 4.3.4 Comparing organism-level and tissue-weighted scores

For comparisons between organism-level and tissue-combined scores we again use a bootstrap approach: for a specific disease term we use the Wilcoxon signed-ranks tests as discussed above to compare performance measures from organism-level scores with tissue-weighted scores. We note that this approach does not take into account: ***(a)*** Variability in the organism-level scores originating form variability of the data they are derived from, and ***(b)*** The possibility that organism-level scores may have already used SNPs in their score derivation process, and we use them again for evaluation in their score derivation process. However, we don’t expect these issue to substantially confound or results, and we note that incurred bias in our comparisons would expected to be in favor of organism-level scores. Results are summarized in **Suppl. Data** SD6, SD7, SD10 and SD11.

#### 4.3.5 DIVAN performance assessment and comparison

To assess and compare our performance with DIVAN [19], we generated a test set of SNPs from the GWAS catalog that were *i)* added after DIVAN had been published (i.e., after 05/28/2016) and *ii)* not present in the database used to train DIVAN (Association Result Browser https://www.ncbi.nlm.nih.gov/projects/gapplus/sgap_plus.htm) and *iii)* not within 1kb distance around SNPs used to train DIVAN and *iv)* were annotated to a disease phenotype addressed by DIVAN.

Control SNPs were generated using SNPsnap matching, as described above. To be able to satisfy criterion *iv)*, we mapped our disease terms (EFO terms) to disease terms used by DIVAN (MeSH terms) using the EMBL-EBI Ontology Xref Service (OxO, https://www.ebi.ac.uk/spot/oxo/, retrieved on April 19, 2020) and were able to resolve 41 out of 45 terms (**Suppl. Data** SD12). Of these, we keep terms with 20 or more disease associated SNPs in the test set and 50 or more SNPs in a training set that we also construct (see below), yielding 29 overall disease phenotypes we use in our analysis. In order to fairly compare DIVAN with our logistic regression approach we constructed a training set using disease-associated SNPs from the GWAS catalog and the Phenotype-Genotype Integrator (PheGenI, https://www.ncbi.nlm.nih.gov/gap/phegeni) [54], excluding SNPs in the test dataset describe above, or SNPs within 1kb around test SNPs. **Suppl. Data** SD13 summarizes test and training data used for this analysis. Results are summarized in **Suppl. Data** SD14.

#### 4.3.6 Performance assessment using chromosome hold-out

To assess the performance of our DHS tissue-weighted score we also used a chromosome hold-out strategy, with test SNPs on different chromosomes from training data. Specifically, for each disease, we choose a set of chromosomes that contains approximately 20% SNVs with a 1/10 positive to negative ratio (the same as the cross-validation setting) as a test set. Selection of test chromosomes is performed for each disease term separately, as disease-associated SNPs differ. To automate the procedure, we deployed (binary) linear programming to pick out chromosomes in test set for each disease.

Specifically, for each disease term we solve the optimization problem

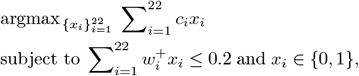

where {*x*_*i*_} are binary indicator variables whether a chromosome is included in the test/hold-out set; 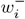 and 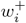 are the fraction of disease-associated 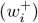 and control SNPs 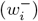 on chromosome *i* and weights in the objective function are defined as 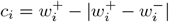. This approach selects, for each disease term, a set of chromosomes to hold out that contain about 20% of disease-associates SNPs and that approximately reflects the overall imbalance between disease-associated and control SNPs. **Suppl. Fig**. S17 and S18 contain performance evaluations on chromosome hold-out sets.

#### 4.3.7 Performance assessment using one SNP per LD block

To assess the effect of SNP correlation on our results we also performed analyses using only a single representative SNP per LD block (defined by *r*^2^ ≥ 0.5, see **Section** 4.1.1). Results are shown in **Suppl. Fig**. S19 and S20.

### 4.4 Comparison with genetic correlation

We retrieved genetic correlation values from the GWAS atlas [31]. To be able to use these data we mapped EFO disease terms (used in the NIH-NCBI GWAS Catalog and in our study) to terms used in the GWAS atlas study. To do so, we extracted synonyms of each EFO term (as listed on EFO ontology) and compared each synonym to the “trait” and “uniqtrait” column in the GWAS atlas data. All matches (with one tolerated letter substitution) will be used.

In this approache a single EFO term can map to multiple GWAS atlas traits and studies. To estimate the genetic correlation between two EFO terms (say *d*_*i*_ and *d*_*j*_), we use a weighted combination of genetic correlation values:

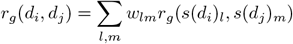

where *r*_*g*_(·, ·) is the genetic correlation of two diseases, 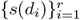 and 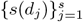 are the GWAS atlas studies that are mapped to EFO term *d*_*i*_ and *d*_*j*_, respectively; *w*_*lm*_ is a weight for each combination of the GWAS atlas studies accounting for the sample sizes of different studies used to estimate genetic correlation values. We choose

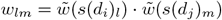

where

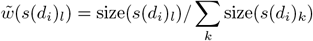

where “size” denotes the sample size of a study. This schene puts higher weights on studies with large sample sizes and smaller weights to studies with smaller sample sizes.

### 4.5 Notes about epimap comparison, cluster annotation and display

#### 4.5.1 Epimap trait-tissue association for Table 5

We obtained the latest snp-centric GWAS enrichments table from the EpiMap Repository at http:////compbio.mit.edu/epimap/. We retrieve tissues with adjusted p-values for each disease. We map the tissue names used in our study (Standard Roadmap Epigenomes, as labed by EID) to tissue names used in epimap (biosamples, as labeld as BSS biosample id) by adapting the scripts from https://github.com/cboix/EPIMAP_ANALYSIS/blob/master/metadata_scripts/get_roadmap_mapping.R. If there are more than one biosamples tissues mapped to roadmap tissues, we reported the p value of the tissue with the most significant results.

#### 4.5.2 Cluster names in Table 6

To name each cluster/group of diseases/EFO terms we choose the EFO term that contains most of the cluster/group members. In **Suppl. Data** SD21 we summarize the terms with high term frequency in each cluster, where term frequency is the fraction of *descendant* terms present. For example, the EFO term “immune system disease” (EFO:0000540) has a term frequency of 0.588 in the “immune-1 cluster”; this means that 58.8% of EFO terms in that cluster are descendants of EFO:0000540. We exclude the terms that are overly broad such as the term “disease” or “experimental factor ontology”. For each cluster, we rank the cluster member EFO terms using term frequency and select as name a meaningful term with the high term frequency. For one cluster where no term had high frequency we chose the name “heterogeneous”.

We also show a diagrams of EFO disease term relationships in each cluster in **Suppl. Fig**. S10-S16. Occasionally we include ancestor EFO terms not present in the cluster in a diagram, which are marked by asterisks.

#### 4.5.3 Dimension reduction and coefficient heatmap

##### UMAP plot

The two-dimensional UMAP plot of 111 EFO disease terms in **Fig**. 7 is based on disease similarities based on context-weighted score averaging (see section 4.2.2). The `umap` function of the `uwot R` package was used with parameters `n_neighbors = 15`, `ret_model = TRUE`, `PCA_center = FALSE`.

##### Coefficient heatmap

The heatmap in **Fig**. 8 displays coefficient vectors of models for disease association (see section 4.2.1), normalized for each disease. Specifically, for each disease and tissue coefficient *x*_*i*_

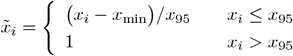

where *x*_min_ is the minimum coefficient for a disease, and *x*_95_ is the 95% quantile.

##### Cluster-associated tissues

For each cluster, we show the top-five tissues that are most associated with the cluster (**Fig**. 8). To identify these tissues we conduct a two-sample Wilcoxon test (one-sided) on every tissue, where we compare normalized tissue coefficients for this cluster to the the other with the highest coefficients on average. The five tissues with the smallest p-value are then selected as top-five tissues.

##### Tissue-associated clusters

For the heatmap with all tissues in **Suppl. Fig**. S9, we assigned a cluster to each tissue. For each tissue, we calculated the median (across disease terms of a cluster) of the normalized coefficients for all clusters; the cluster with the highest median was assigned.

## Data Availability

All data produced in the present study are available upon reasonable request to the authors.

## Data and code availability

Public data repositories were used as detailed in the Methods section, and data underlying tables and figures is available as supplemental information online. 25bp-resolution tissue-weighted DHS scores are available for download at https://doi.org/10.7910/DVN/AUAJ7K, and computer code used to generate analyses presented is available at link-to-github.

## Notes

### Competing Interest Statement

The authors have declared no competing interest.

### Funding Statement

This study did not receive any funding

### Author Declarations

This study used only openly available human data that were located at the URLs provided in the methods section.

