## Supplemental figures and tables for "Disease-specific prioritization of non-coding GWAS variants based on chromatin accessibility"

### Supplementary Information

...title...

### 1 Supplemental Methods

#### Four different matching strategies for control SNVs

For each disease-associated SNV, we have matched control SNVs from four different matching strategies: 1) random; 2) SNPsnap_TSS; 3) SNPsnap; and 4) TSS (see **Method**). We measured the performance of five organism level scores on four different control sets in 111 diseases. Among them, random matching is considered as the least stringent way as we don't have any constraint on it. Therefore, we choose the random matching as the baseline, and we normalize the performance of the other three control sets on random matching for each disease. We plot the normalized performance in **Suppl. Fig.** S1 and S2, using each disease as a panel.

From here, we observe that three normalized performances in CADD are all distributed around 1. This indicates that the CADD is robust in different matching strategies. The normalized performances of Eigen, GenoCanyon, GWAVA and LINSIGHT are all less than 1. This indicates that those three matched control sets are more stringent than randomly selecting control variants. Among those three control sets, TSS is the most stringent, followed by SNPsnap and SNPsnap_TSS.

It is important to note that TSS and SNPsnap TSS are both matched using the distance to the nearest TSS; however, TSS uses the distance to the nearest protein-coding gene while SNPsnap_TSS uses the distance to the nearest gene. TSS matched SNVs have similar distribution to disease SNVs in both all genes and protein-coding genes; in contrast, SNPsnap_TSS SNVs have similar distribution to disease SNVs in all genes but not in protein-coding genes (**Suppl. Fig.** S3 and S4). Therefore, TSS is more stringent than SNPsnap_TSS and also more stringent than SNPsnap even though SNPsnap has matched with additional three criteria.

Here, SNPsnap matching strategy is neither too stringent nor too loose and it matches with four criteria (see **Method**). Thus, we choose the control set matched by SNPsnap in our study.

#### DHS-weighted performance using two additional strategies to prevent overfitting

To prevent overfitting, we also deployed two additional strategies to test the performance of tissue-weighted DHS. In the first one, we used the 'representative SNVs' so that any two disease-associated SNVs are not in the same LD block. In the second one, we deployed a chromosome held-out strategy so that the SNVs in the test and train set are on different chromosomes (see **Method**). These two strategies ensure that the SNVs in the test and train set are seperated or in different chromosome to reduce overfitting. We observe that in any of these two settings, we can still observe a significant increase with the tissue-weighted model, which is consistent with our previous finding, even though the amount of the improvement is in a lesser degree in some diseases (See See **Suppl. Fig.** S17-S20).

### 2 Supplemental Tables

| Score | Wins | Losses | Ties | Wins (agg) | Losses (agg) | Ties (agg) |
|---|---|---|---|---|---|---|
| GenoCanyon | 335 | 87 | 22 | 4 | 0 | 0 |
| LINSIGHT | 303 | 124 | 17 | 3 | 1 | 0 |
| eigen | 247 | 177 | 20 | 2 | 2 | 0 |
| GWAVA | 168 | 256 | 20 | 1 | 3 | 0 |
| CADD | 15 | 424 | 5 | 0 | 4 | 0 |

Supplemental Table ST1: ***Relative performance of organism-level variant scores, measured by AUROC***. Wins, Losses, Ties refers to significantly better (or worse, or tied) performance across all possible pairings (see **Methods**). The first three columns summarize separate comparisons for each disease term (for each row there are four other methods and 111 terms), while the last three columns represent results of aggregate comparisons across terms. Average precision was used as the performance metric, and the Wilcoxon singed-ranks test to determine wins and losses (p-values less than 0.05 were ties).

| Score | Wins | Losses | Ties | Wins (agg) | Losses (agg) | Ties (agg) |
|---|---|---|---|---|---|---|
| DHS | 138 | 54 | 30 | 2 | 10 | 0 |
| Fitcons2 | 80 | 111 | 31 | 0 | 1 | 1 |
| Genoskyline | 69 | 122 | 31 | 0 | 1 | 1 |

Supplemental Table ST2: ***Relative performance of disease-specific (tissue-weighted) variant scores, measured by AUROC***. Wins, Losses, Ties refers to significantly better (or worse, or tied) performance across all possible pairings (see **Methods**). The first three columns summarize separate comparisons for each disease term (for each row there are two other methods and 111 terms), while the last three columns represent results of aggregate comparisons across terms. Average precision was used as the performance metric, and the Wilcoxon singed-ranks test to determine wins and losses (p-values less than 0.05 were ties).

| Score/Method | By disease term | | | Aggregated | | |
|---|---|---|---|---|---|---|
| | Wins | Losses | Ties | Wins | Losses | Ties |
| DHS | 375 | 127 | 53 | 4 | 0 | 1 |
| GenoCanyon | 375 | 144 | 36 | 4 | 0 | 1 |
| LINSIGHT | 342 | 184 | 29 | 3 | 2 | 0 |
| eigen | 273 | 250 | 32 | 2 | 3 | 0 |
| GWAVA | 186 | 338 | 31 | 1 | 4 | 0 |
| CADD | 19 | 527 | 9 | 0 | 5 | 0 |

Supplemental Table ST3: ***DHS outperforms organism-level variant scores, measured by AUROC***. Wins, Losses, Ties refer to significantly better (or worse, or tied) performance across all possible score pairings (see **Methods**). The first three columns summarize separate comparisons for each disease term (for each row there are two other methods and 111 terms, i.e., 555 comparisons), while the last three columns represent results of comparisons aggregated over terms. Average precision was used as the performance metric, and the Wilcoxon singed-ranks test to determine wins and losses (p-values less than 0.05 were reported as ties).

| Score/Method | Performance vs. DHS | | | |
| --- | --- | --- | --- | --- |
| | Wins | Losses | Ties | Winning percent |
| LINSIGHT | 17 | 84 | 10 | 20 |
| GenoCanyon | 7 | 92 | 12 | 12 |
| GWAVA | 12 | 93 | 6 | 14 |
| eigen | 4 | 98 | 8 | 8 |
| CADD | 4 | 107 | 2 | 4 |

Supplemental Table ST4: ***Disease-specific variant prioritization outperforms organism-level approaches, measured by AUPR***. Wins losses and ties of organism-level scores against tissue-weighted DHS scores (performance measured by average precision, Wilcoxon signed-ranks test for determining significance). Winning percent was calculated as number of wins plus half the number of ties, divided by the number of comparisons, and rounded to the nearest integer. Rows have been ordered by winning percent.

| Score/Method | Performance vs. DHS | | | |
| --- | --- | --- | --- | --- |
| | Wins | Losses | Ties | Winning percent |
| GenoCanyon | 40 | 57 | 14 | 36 |
| LINSIGHT | 39 | 60 | 12 | 35 |
| eigen | 26 | 73 | 12 | 23 |
| GWAVA | 18 | 82 | 11 | 16 |
| CADD | 4 | 103 | 4 | 4 |

Supplemental Table ST5: ***Disease-specific variant prioritization outperforms organism-level approaches, measured by AUROC***. Wins losses and ties of organism-level scores against tissue-weighted DHS scores (performance measured by average precision, Wilcoxon signed-ranks test for determining significance). Winning percent was calculated as number of wins plus half the number of ties, divided by the number of comparisons, and rounded to the nearest integer. Rows have been ordered by winning percent.

| Score | Wins | Losses | Ties | Winning percent |
| --- | --- | --- | --- | --- |
| GenoCanyon | 31 | 25 | 2 | 55 |
| DHS | 27 | 26 | 5 | 51 |
| DIVAN | 24 | 31 | 3 | 44 |

Supplemental Table ST6: ***DHS tissue-weighted disease-specific scoring outperforms DIVAN***. Across 30 disease terms, this table summarizes all pairwise comparison for DHS tissue-weighted, GenoCanyon and DIVAN using a specifically created test dataset. Wins, losses, ties refer to significantly better (or worse, or tied) performance. Average precision was used as the performance metric, and the Wilcoxon singed-ranks test to determine wins and losses (p-values less than 0.05 were ties). Winning percent = #Wins/(#Wins+#Losses)

| Rank | ID | Tissue name | Group | epimap FDR |
|---|---|---|---|---|
| Systemic scleroderma | | | | |
| 1 | E116 | GM12878 Lymphoblastoid Cells | blood | 0.04 |
| 2 | E032 | Primary B cells from peripheral blood | blood | 0.16 |
| 3 | E041 | Primary T helper cells PMA-I stimulated | blood | 0.08 |
| 4 | E123 | K562 Leukemia Cells | blood | 1.00 |
| 5 | E030 | Primary neutrophils from peripheral blood | blood | 0.86 |
| Sclerosing cholangitis | | | | |
| 1 | E116 | GM12878 Lymphoblastoid Cells | blood | <0.001 |
| 2 | E061 | Foreskin Melanocyte Primary Cells skin03 | skin | 0.18 |
| 3 | E102 | Rectal Mucosa Donor 31 | gi_rectum | <0.001 |
| 4 | E041 | Primary T helper cells PMA-I stimulated | blood | <0.001 |
| 5 | E029 | Primary monocytes from peripheral blood | blood | <0.001 |
| Colorectal adenoma | | | | |
| 1 | E102 | Rectal Mucosa Donor 31 | gi_rectum | 0.05 |
| 2 | E110 | Stomach Mucosa | gi_stomach | 0.008 |
| 3 | E057 | Foreskin Keratinocyte Primary Cells skin02 | skin | 0.11 |
| 4 | E101 | Rectal Mucosa Donor 29 | gi_rectum | 0.003 |
| 5 | E028 | Breast variant Human Mammary Epithelial Cells (vHMEC) | breast | 0.20 |
| Atrial fibrillation | | | | |
| 1 | E083 | Fetal Heart | heart | <0.001 |
| 2 | E108 | Skeletal Muscle Female | muscle | 0.01 |
| 3 | E107 | Skeletal Muscle Male | muscle | 0.008 |
| 4 | E088 | Fetal Lung | lung | 0.002 |
| 5 | E120 | HSMM Skeletal Muscle Myoblasts Cells | muscle | 0.18 |
| Cutaneous melanoma | | | | |
| 1 | E061 | Foreskin Melanocyte Primary Cells skin03 | skin | 0.08 |
| 2 | E059 | Foreskin Melanocyte Primary Cells skin01 | skin | 0.08 |
| 3 | E117 | HeLa-S3 Cervical Carcinoma Cell Line | cervix | 0.39 |
| 4 | E041 | Primary T helper cells PMA-I stimulated | blood | 0.49 |
| 5 | E122 | HUVEC Umbilical Vein Endothelial Primary Cells | vascular | 0.88 |

Supplemental Table ST7: ***Top-ranked tissues for five diseases.*** For five diseases when show the top-five tissues with the largest tissue weights in the corresponding model we derive. The first column is the tissue rank, the second the tissue's roadmap ID, the third the tissue name, the fourth the tissue group, and the fifth listst the adjusted p-value in an enrichment analysis performed by epimap [**?**].

### 3 Supplemental Data Legends

#### SD1 Phenotypes used in this study

`Filename: sup_data_disease-terms.csv.gz`

The first column denotes the EFO name of disease phenotypes used. Column #2 is the EFO ID. Column #3 shows the number of SNVs associated with the term (coding and non-coding). Columns #4 shows the number of non-coding SNVS used in the study before aggregation and #5 shows the number of non-coding SNVs used after aggregation. Non-EUR 1KG SNVs and SNVs in the HLA region have been removed in column #4 and #5.

#### SD2 Disease-associated SNVs used in this study

`Filename: sup_data_disease-snvs.csv.gz`

The first column denotes the SNV ID. Column #2 is the rsID. Column #3 is the phenotype. Columns #4 and 5 are the chromosome and the specific location (hg19 coordinates). Column #6 is the LD block cluster id where this SNV resides(SNVs in the same LD block will have the same cluster id), and column #7 indicates whether this SNV is selected as the representative SNV for the block (1 as selected, 0 as not selected). SNVs associated with multiple diseases appear in more than one row.

#### SD3 Control SNVs used in this study

`Filename: sup_data_control-snvs.csv.gz`

For each disease-associated SNV, this table lists ∼10 randomly-selected control SNVs by four different methods (see **Methods**). The first column denotes the SNV ID. Column #2 is the rs ID. Column #3 is the phenotype. Column #4 and 5 are the chromosome and the specific location (hg19) of that SNV. Column #6 is the matching strategy (i.e. snpsnap, snpsnap_tss, tss, random) and column #7 is the SNV ID of the corresponding disease-associated SNV.

#### SD4 Pairwise comparisons of organism-level scores for each disease term

`Filename: sup_data_pairwise-org-individual.csv.gz`

For each combination of organism-level scores we report p-values for a Wilcoxon signed-ranks test for each individual disease (see **Methods**). Column #1 is the score name. Column #2 is the median performance across bootstrap runs for that score. Column #3 is the second score name. Column #4 is the median performance for the second score. Column #5 is the disease term for which the comparison was performed. Column #6 is the curve type we used for the area under the curve performance metric (ROC or PR). Column #7 is the p-value of the test. Column #8 is the score with the higher median.

#### SD5 Pairwise comparisons of organism-level scores, aggregated across diseases

`Filename: sup_data_pairwise-org-aggregated.csv.gz`

For each combination of organism-level scores we report p-values for a Wilcoxon signed-ranks test, aggregated across 111 diseases (see **Methods**). Column #1 is the score name. Column #2 is the median performance across 111 diseases. Column #3 is the second score name. Column #4 is the median performance for the second score across 111 diseases. Column #5 is the curve type for the area under the curve performance metric (ROC or PR). Column #6 is the p-value of the test. Column #7 is the score with the higher median.

#### SD6 Pairwise comparison of Tissue-weighted scores vs. Tissue-mean scores for each disease term

`Filename: sup_data_pairwise-tis-individual.csv.gz`

For each score we report p-values for a Wilcoxon signed-ranks test between the tissue-mean and tissue-weighted version for each individual disease. Column #1 is the score name. Column #2 is the median performance across bootstrap runs for that score. Column #3 is the second score name. Column #4 is the median performance for the second score. Column #5 is the disease term for which the comparison was performed. Column #6 is the curve type we used for the area under the curve performance metric (ROC or average precision). Column #7 is the p-value of the test. Column #8 is the score with the higher median.

#### SD7 Pairwise comparison of Tissue-weighted scores vs. Tissue-mean scores, aggregated across diseases

`Filename: sup_data_pairwise-tis-aggregated.csv.gz`

For each score we report p-values for a Wilcoxon signed-ranks test between the tissue-mean and tissue-weighted version, aggregated across all diseases. Column #1 is the score name. Column #2 is the median performance across all diseases for that score. Column #3 is the second score name. Column #4 is the median performance for the second score. Column #5 is the curve type for the area under the curve performance metric (ROC or PR). Column #6 the p-value of the test. Column #7 is the score with the higher median.

#### SD8 Pairwise comparison of three Tissue-weighted scores for each disease term

`Filename: sup_data_pairwise-tis-weighted-individual.csv.gz`

For each combination of Tissue-weighted scores we report p-values for a Wilcoxon signed-ranks test for each individual disease (see **Methods**). Column #1 is the score name. Column #2 is the median performance across bootstrap runs for that score. Column #3 is the second score name. Column #4 is the median performance for the second score. Column #5 is the disease term for which the comparison was performed. Column #6 is the curve type we used for the area under the curve performance metric (ROC or PR). Column #7 is the p-value of the test. Column #8 is the score with the higher median.

#### SD9 Pairwise comparison of three Tissue-weighted scores, aggregated across diseases

`Filename: sup_data_pairwise-tis-weighted-aggregated.csv.gz`

For each combination of Tissue-weighted scores we report p-values for a Wilcoxon signed-ranks test, aggregated across all diseases (see **Methods**). Column #1 is the score name. Column #2 is the median performance across all diseases for that score. Column #3 is the second score name. Column #4 is the median performance for the second score. Column #5 is the curve type for the area under the curve performance metric (ROC or PR). Column #6 is the p-value of the test. Column #7 is the score with the higher median.

#### SD10 Pairwise comparison of Tissue-weighted-DHS vs five organism-level scores for each disease term

`Filename: sup_data_pairwise-tis-vs-org-individual.csv.gz`

We report p-values for a Wilcoxon signed-ranks test between the Tissue-weighted-DHS and five organism-level scores for each individual disease. Column #1 is the score name. Column #2 is the median performance across bootstrap runs for that score. Column #3 is the second score name. Column #4 is the median performance for the second score. Column #5 is the disease term for which the comparison was performed. Column #6 is the curve type we used for the area under the curve performance metric (ROC or PR). Column #7 is the p-value of the test. Column #8 is the score with the higher median.

#### SD11 Pairwise comparison of tissue-weighted-DHS and five organism-level scores aggregated

`Filename: sup_data_pairwise-tis-org-aggregated.csv.gz`

We report p-values for a Wilcoxon signed-ranks test between the Tissue-weighted-DHS and five organism-level scores, aggregated across all diseases. Column #1 is the score name. Column #2 is the median performance across bootstrap runs for that score. Column #3 is the second score name. Column #4 is the median performance for the second score. Column #5 is the curve type we used for the area under the curve performance metric (ROC or PR). Column #6 is the p-value of the test. Column #7 is the score with the higher median.

#### SD12 Mapping of mesh terms to EFO terms

`Filename: sup_data_mapping-efo-mesh.csv.gz`

The first and second columns are the mesh term id and mesh term label used by DIVAN. The third and fourth columns are the EFO ID and EFO label that is mapped to the mesh terms. (Note: there are two MeSH terms that are matched to more than 1 EFO term.)

#### SD13 Training and test SNVs used to compare Tissue-weighted with DIVAN (including disease and matched control SNVs)

`Filename: sup_data_divan-snvs.csv.gz`

Column #1 denotes the SNV ID. Column #2 is the rs ID of the SNV. Column #3 is the phenotype. Column #4 and #5 are the chromosome and location of the SNV. Column #6 indicates whether the variant is a disease-associated or a control variant. Column #7 is the SNV ID of the corresponding disease-associated SNV. Column #8 indicates whether the variant is in training or test set.

#### SD14 Pairwise comparison of DIVAN vs. GenoCanyon vs Tissue-weighted-DHS for each disease term

`Filename: sup_data_pairwise-divan-individual.csv.gz`

For each combination of DIVAN vs. GenoCanyon vs. Tissue-weighted-DHS we report p-values for a Wilcoxon signed-ranks test for each individual disease (see **Methods**). Column #1 is the score name. Column #2 is the median performance across bootstrap runs for that score. Column #3 is the second score name. Column #4 is the median performance for the second score. Column #5 is the disease term for which the comparison was performed. Column #6 is the curve type we used for the area under the curve performance metric (ROC or PR). Column #7 is the p-value of the test. Column #8 is the score with the higher median.

#### SD15 GenoCanyon vs DIVAN in our study and in Chen study (the DIVAN study)

`Filename: sup_data_perf-divan-our-vs-chen.csv.gz`

Column #1 is the disease names of 27 overlapping diseases. Column #2 indicates whether GenoCanyon is better than DIVAN in our study. Column #3 indicates whether GenoCanyon is better than DIVAN as published by DIVAN.

#### SD16 Tissue-weighted prediction scores for SNVs across 111 diseases

`Filename: sup_data_prediction-scores-dhs-weighted.csv.gz`

Column #1 is the the SNV_ID (chr:position). (If a SNV is annotated to multiple phenotypes, there will be multiple entries.) Column #2 is the phenotype that is annotated to the SNVs. Column #3 indicates whether this SNV is a disease-associated variant or a control variant. Column #4-6 are Tissue-weighted prediction scores in Genoskyline, DHS and Fitcons2

#### SD17 Beta coefficients of the logistic regression models in 111 diseases (using DHS score)

`Filename: sup_data_beta-coefficients-mean-dhs.csv.gz`

Column #1 is the phenotypes. Column #2-128 are the mean of the coefficients of 127 tissues.

#### SD18 Standard deviation of the beta coefficients in SD1

`Filename: sup_data_beta-coefficients-sd-dhs.csv.gz`

Column #1 is the phenotypes. Column #2-128 are the standard deviation of the coefficients in 127 tissues.

#### SD19 Disease-disease similarities derived from the logistic regression model (DHS)

`Filename: sup_data_beta-model-similarity-dhs.csv.gz`

column #1 and column #2 are the names of the disease pairs. Column #3 is the weighted disease-disease similarity derived from the model.

#### SD20 Clusters assigned to 111 diseases

`Filename: sup_data_cluster-id-name.csv.gz`

Column #1 is the disease name. Column #2 is the cluster id. Column #3 is the cluster name.

#### SD21 Term frequency in 7 disease clusters

`Filename: sup_data_cluster_term_frequency.csv.gz`

Column #1 is the term name. Column #2 is the term id. Column #3 is the term frequency of a term in the cluster. Column #4 is cluster id. Term frequency means the fraction of diseases in this cluster that is a descendant of this term. For example, immune system disease with a term frequency 0.588 in cluster immune-1 means that 58.8% of diseases in immune-1 cluster is a immune system disease.

#### SD22 Top five tissues in 7 disease clusters

`Filename: sup_data_top-five-tissues.csv.gz`

Column #1 and #2 are the cluster id and name. Column #3-5 are the tissue id, tissue name and tissue anatomy.

#### SD23 ID, name and group of standard epigenomes

`Filename: sup_data_standard-epigenomes`

Column #1 is the ID of the standard epigenomes (e.g. E043). Column #2 is the group name (e.g. Blood & T-cell). Column #3 is the standardized epigenome name. Column #4 is the anatomy(e.g. blood). Column #5 is the type (e.g. PrimaryCell).

#### SD24 Genetic correlation of the disease pairs

`Filename: sup_data_genetic-correlation.csv.gz`

column #1 and column #2 are the name of the disease pairs. Column #3 is the genetic correlation derived from the GWAS ATLAS

#### SD25 Performance of Tissue-weighted (DHS) in different held-out strategies

`Filename: sup_data_perf-chrom-heldout.csv.gz`

Column #1 is the disease name. Column #2-#10 are the performance of Tissue-weighted (DHS) and Tissue-mean (DHS) measured in different held-out strategies. CV-B: cross-validation, baseline; CV-LR: cross-validation logistic regression; CV-LR (SD): standard deviation of CV-LR; random-B: randomly sampled test set, baseline; random-B (SD): random-B standard deviation; random-LR: randomly sample test set, logistic regression; random-LR (SD): random-LR standard deviation; chr-B: test set held out by chromosome, baseline; chr-B (SD): chr-B standard deviation.

#### 4 Supplemental Figures

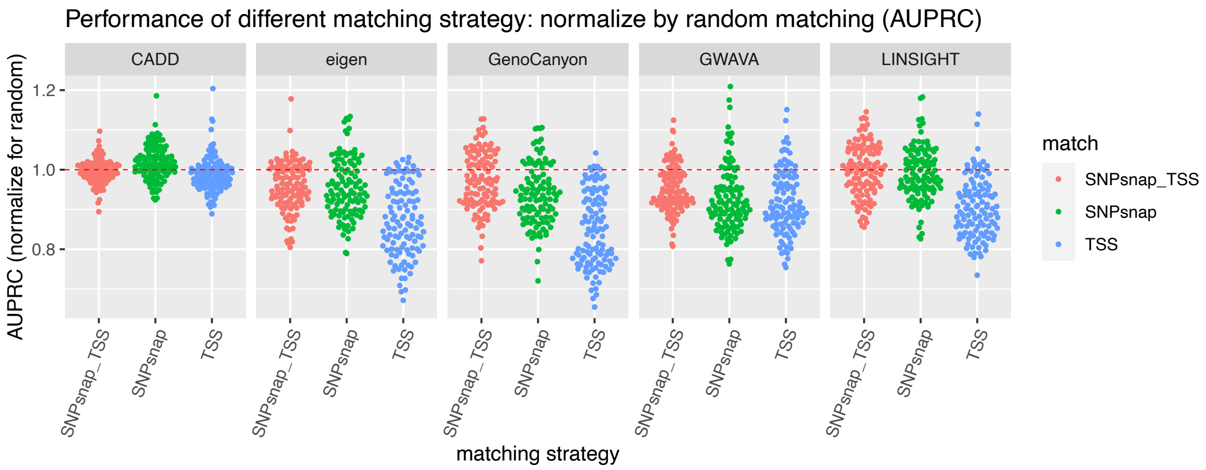


Supplemental Figure S1: ***Performance of different matching strategy, measured by area under the PR curve***. X-axis delineates three different matching strategies (i.e. snpsnap-tss, snpsnap, tss). Y axis shows the performance in terms of area under precision recall curve, normalized by random matching. Each point represents a specific disease term. Horizontal lines spanning the dataset denotes the scenario that the normalized performance equals to 1.

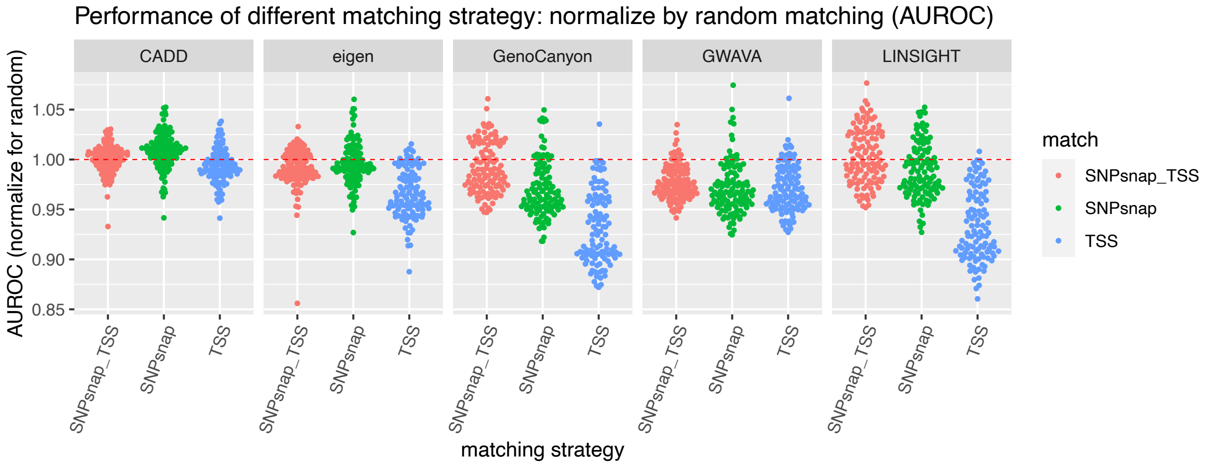


Supplemental Figure S2: ***Performance of different matching strategy, measured by area under the ROC curve***. X-axis delineates three different matching strategies (i.e. snpsnap-tss, snpsnap, tss). Y axis shows the performance in terms of area under receiver operating characteristic curve, normalized by random matching. Each point represents a specific disease term. Horizontal lines spanning the dataset denotes the scenario that the normalized performance equals to 1.

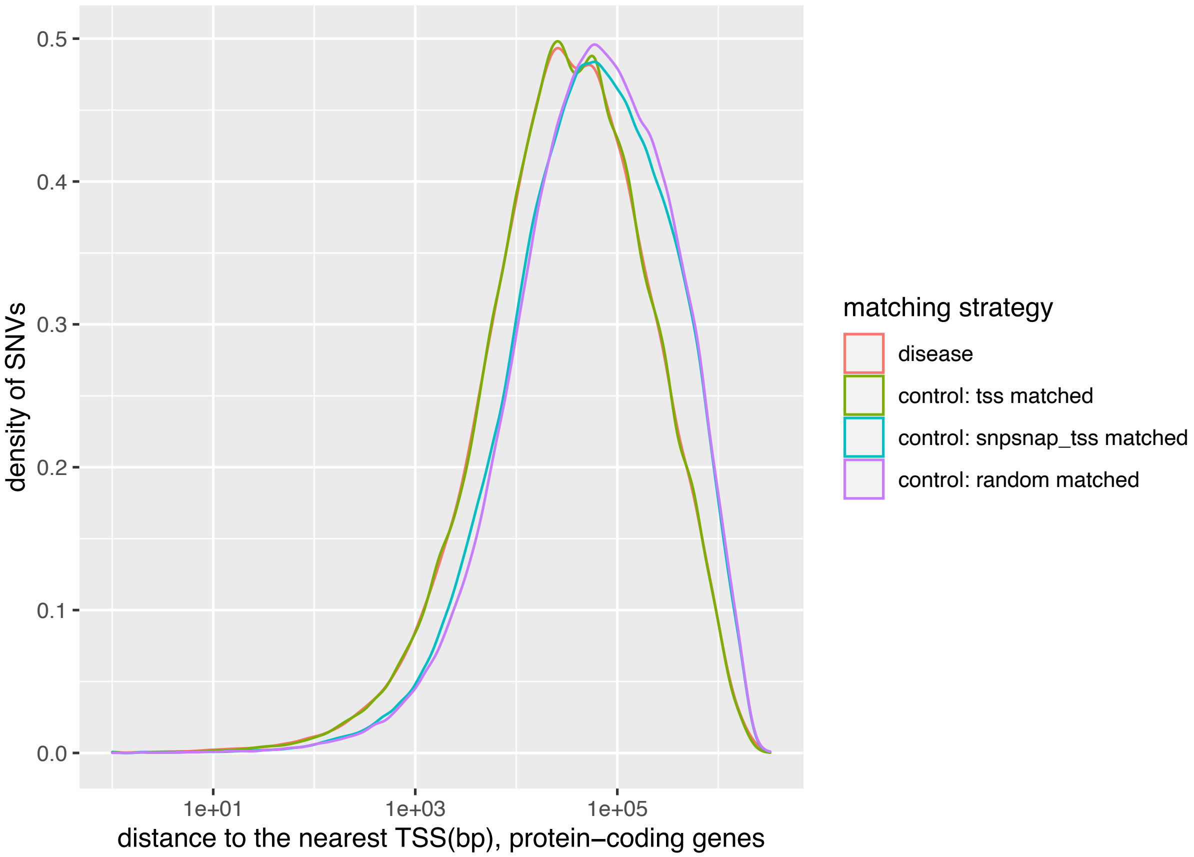


Supplemental Figure S3: ***A density plot showing the distribution of distance to nearest TSS (protein coding genes) in disease SNVs and three different control SNVs***. X-axis shows the distance to the nearest TSS of the protein-coding genes and is log 10 scaled. Y axis shows the density of SNVs.

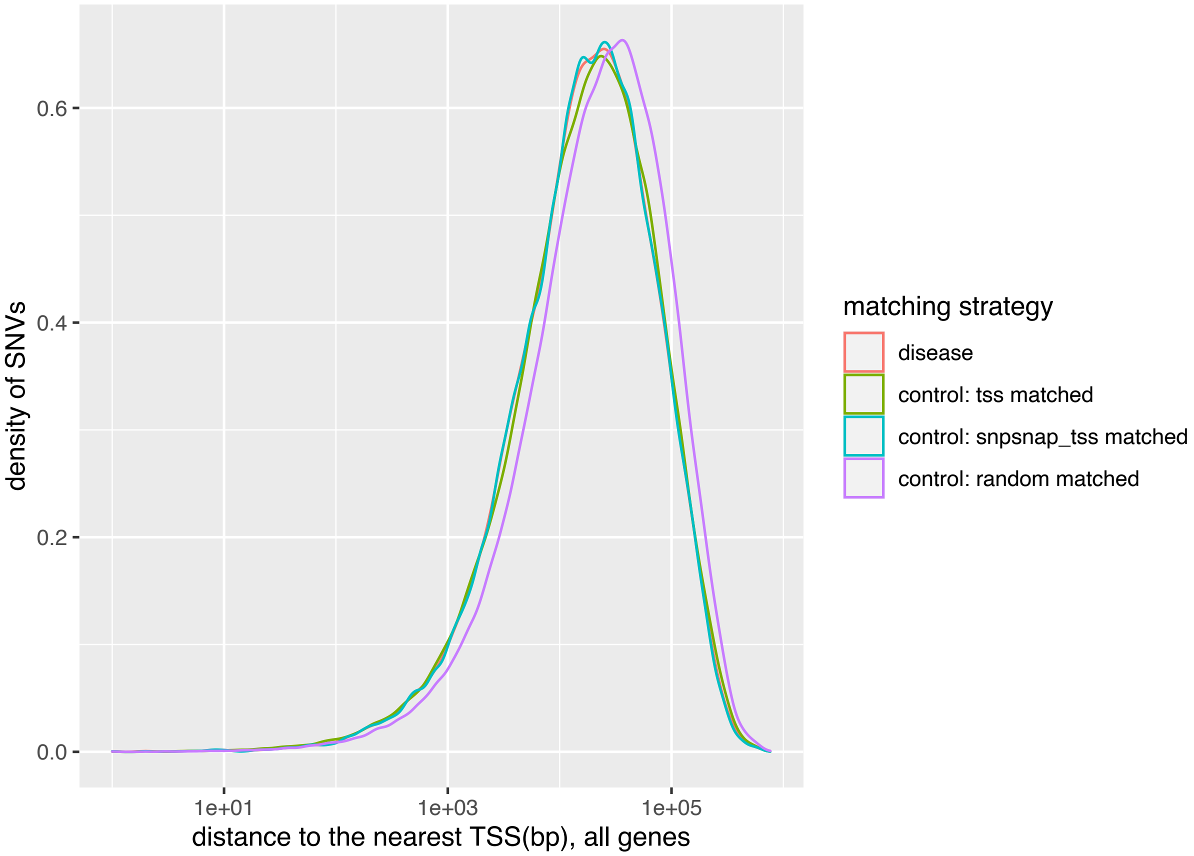


Supplemental Figure S4: ***A density plot showing the distribution of distance to nearest TSS (all genes) in disease SNVs and three different control SNVs***. X-axis shows the distance to the nearest TSS of the protein-coding genes and is log 10 scaled. Y axis shows the density of SNVs.

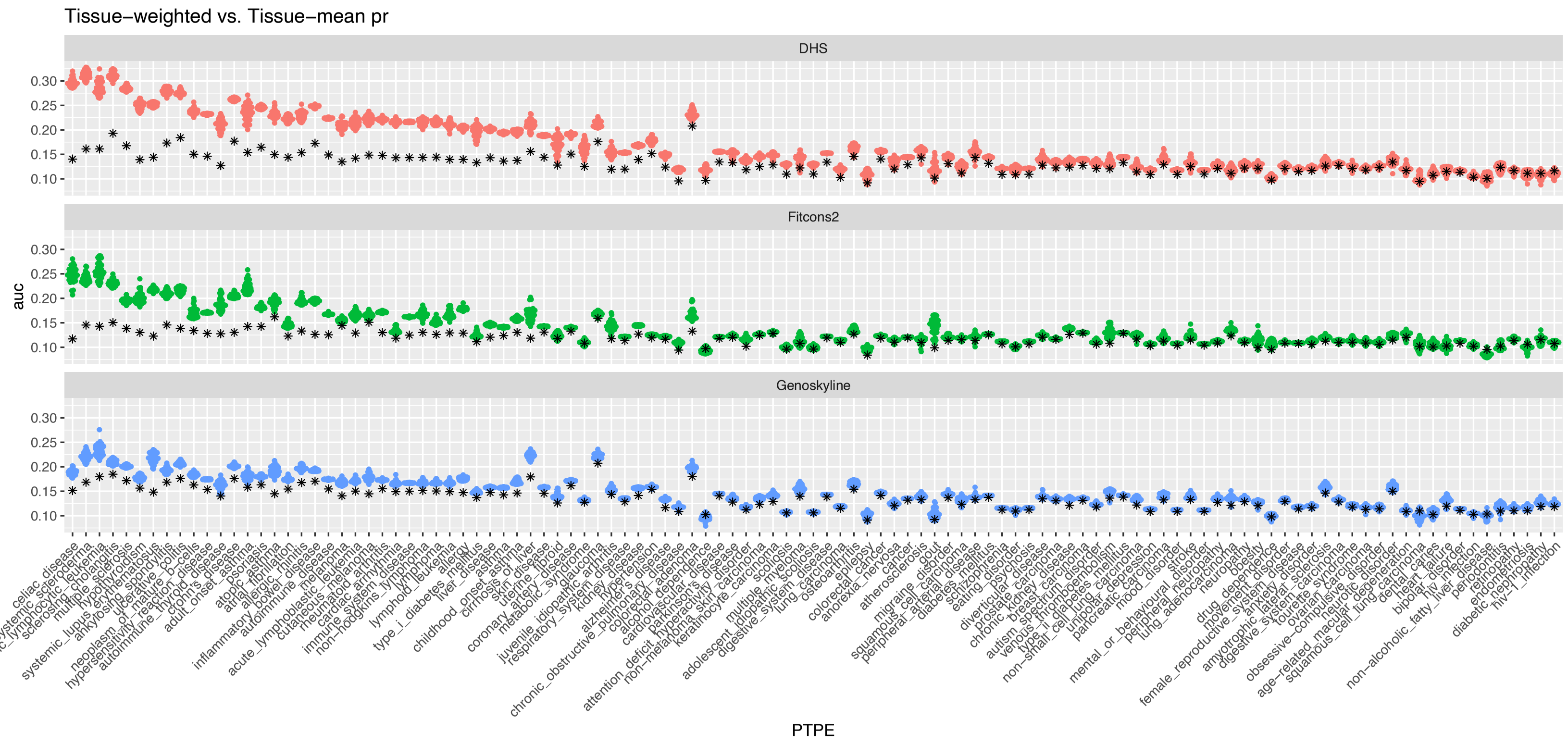


Supplemental Figure S5: ***Tissue-Weighted performance compared with Tissue-Mean in 111 diseases***. X-axis delineates different diseases and y-axis is the performance in terms of area under the precision recall curve. The star represents the Tissue-Mean and the colored dots are Tissue-Weighted with 30 replicates.

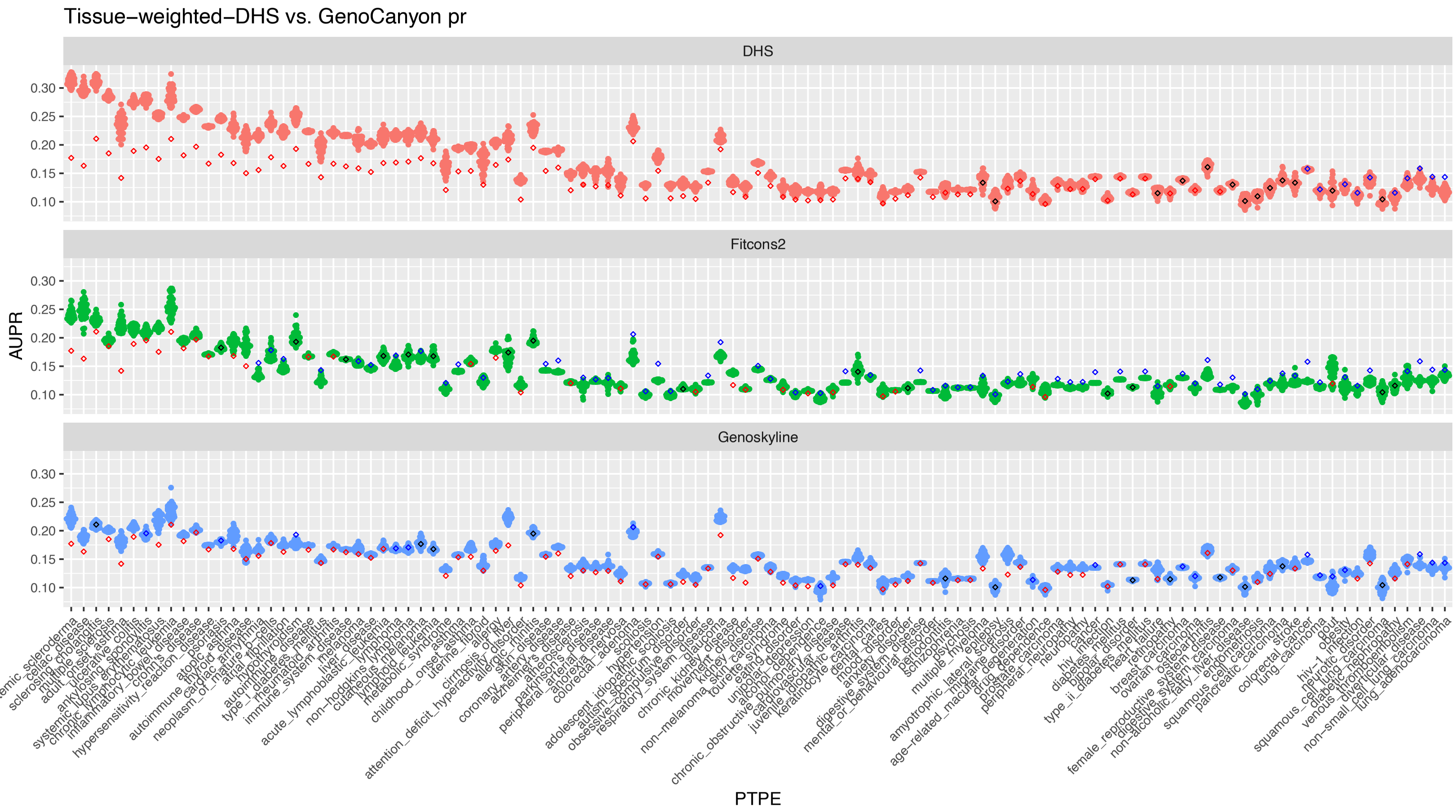


Supplemental Figure S6: ***Tissue-Weighted-DHS performance compared with GenoCanyon in 111 diseases***. X-axis delineates different diseases and y-axis is the performance in terms of area under the precision recall curve. The diamond represents the GenoCanyon (red: Tissue-Weighted better; black: comparable performance; blue: GenoCanyon better) and the colored dots are Tissue-Weighted with 30 replicates.

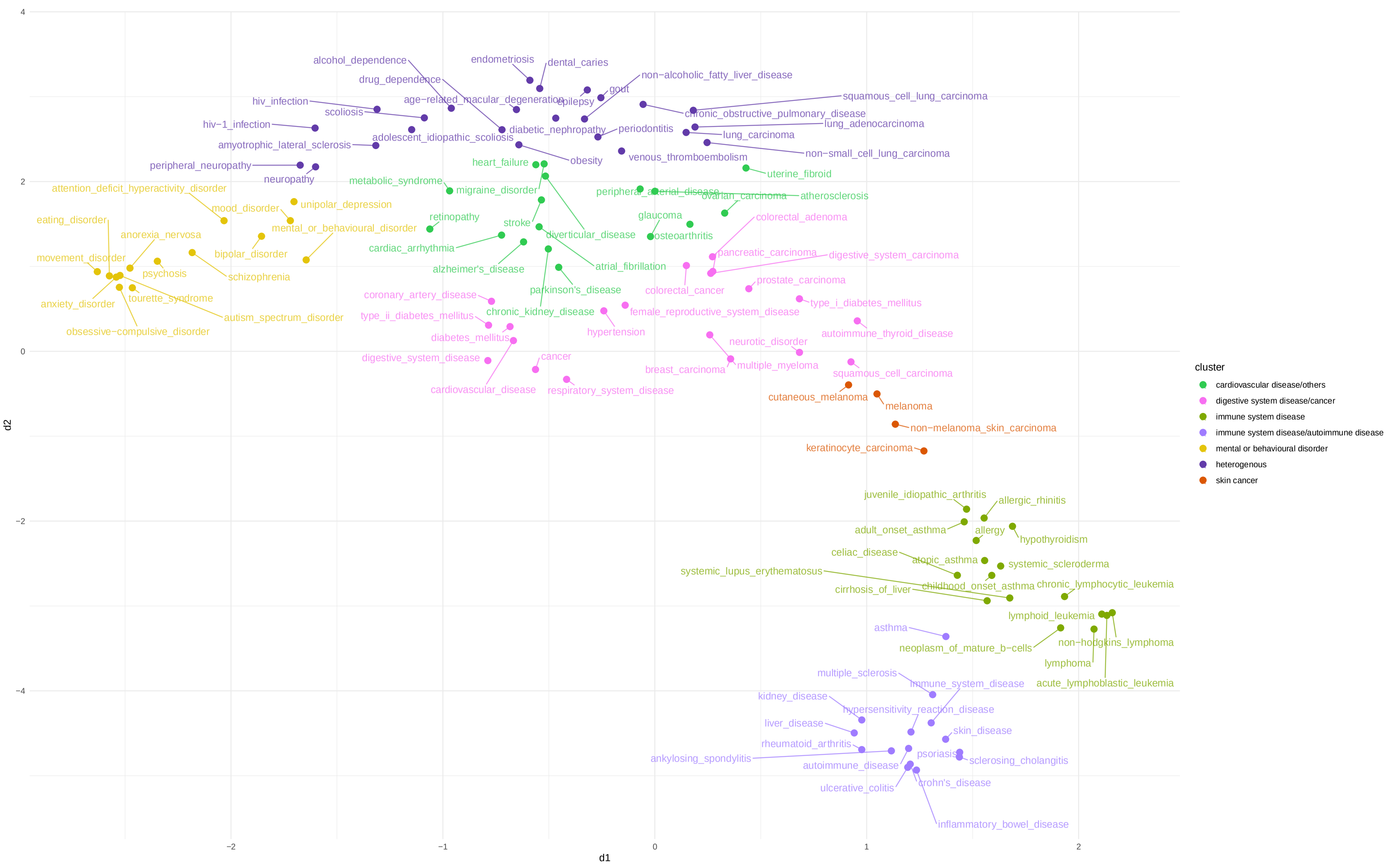


Supplemental Figure S7: ***Umap plot shows 7 clusters of 111 diseases***. Hierarchical clustering was used to group diseases into 7 clusters.

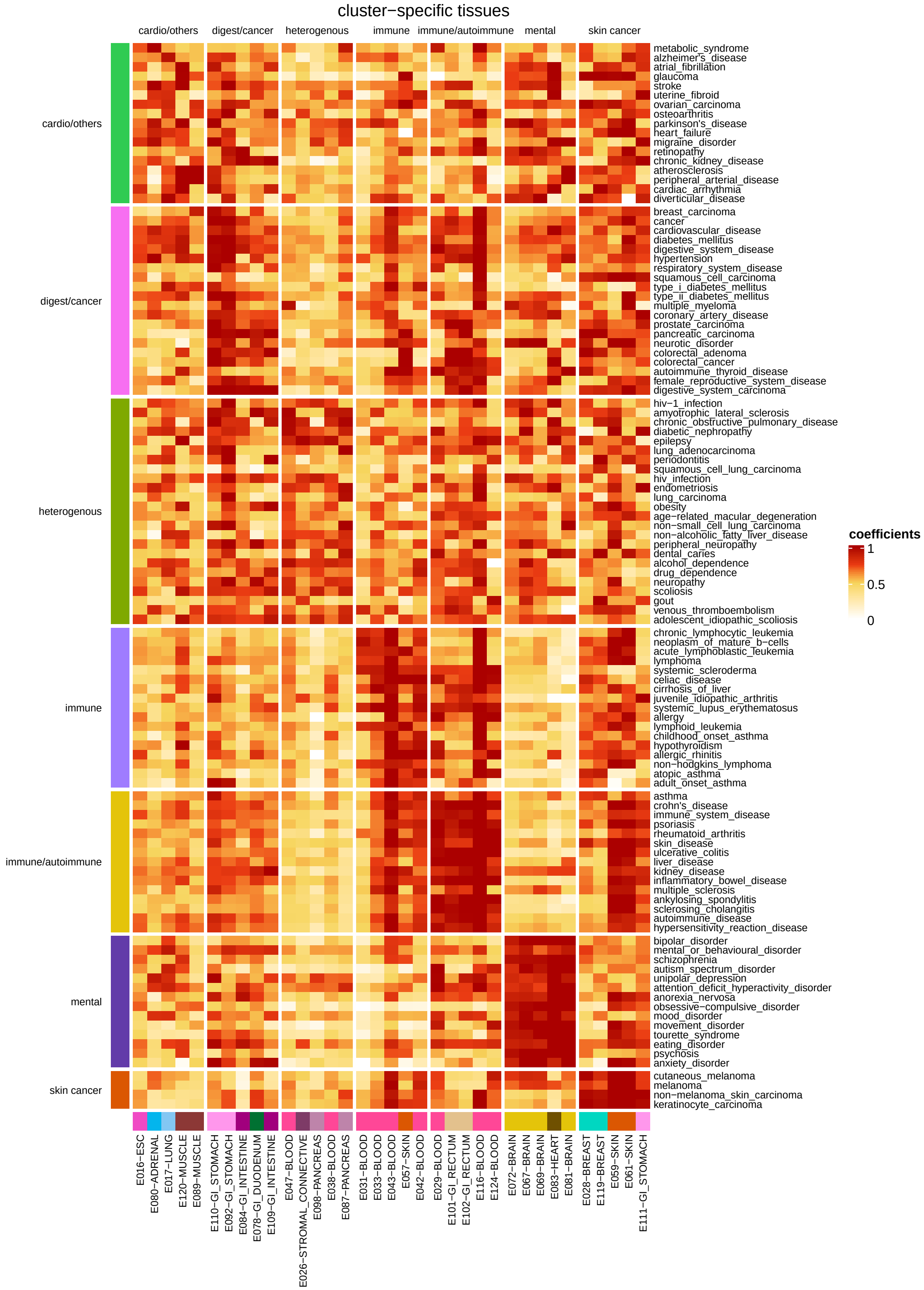


Supplemental Figure S8: ***Heatmap plot of coefficients of 111 diseases*** . Coefficients are regularized by each disease.

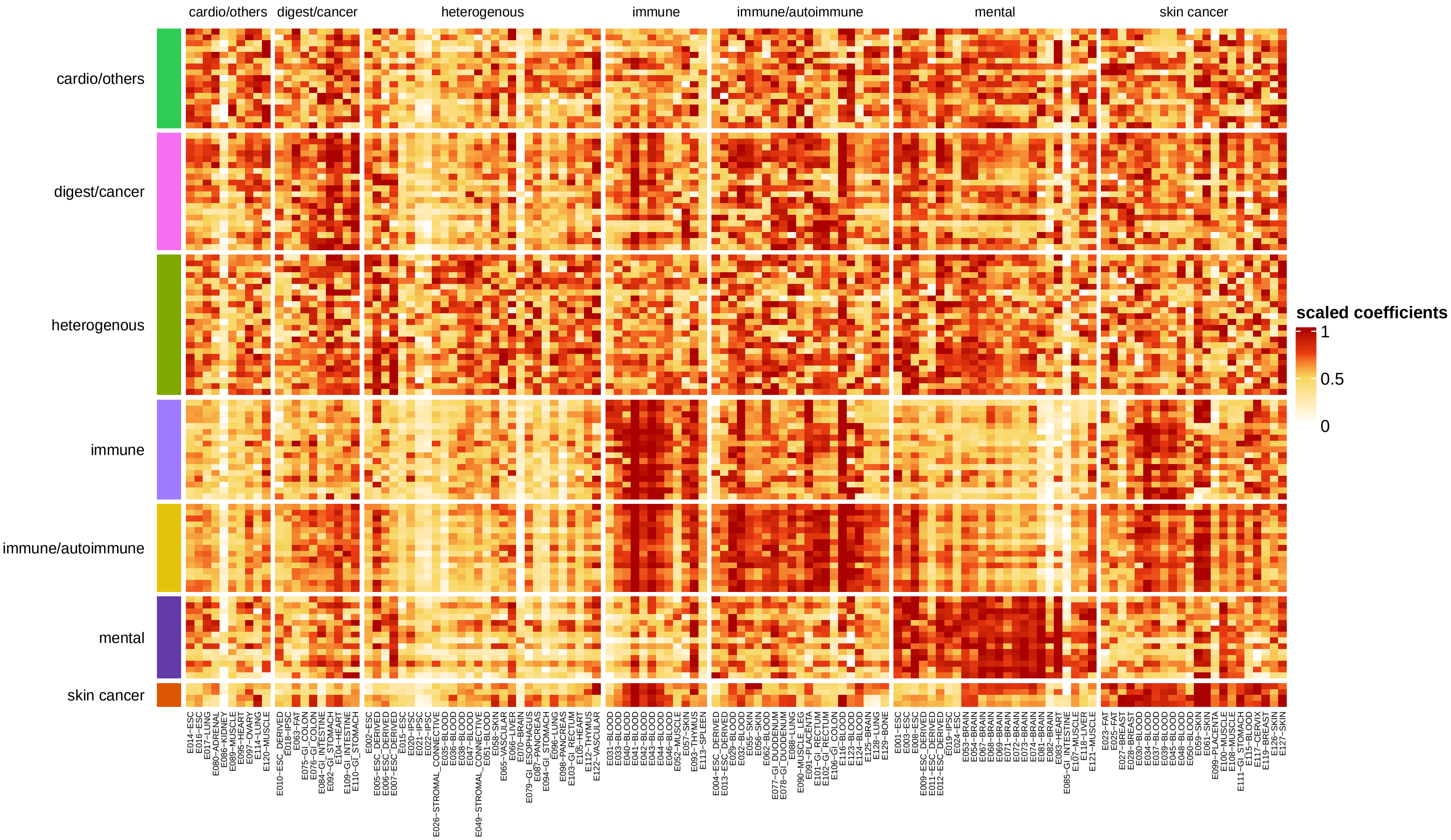


Supplemental Figure S9: ***Heatmap plot of coefficients of 111 diseases on 5 cluster-specific tissues***. Coefficients are regularized by each disease. Tissue names are shown by 'Tissue name-group' from 127 standard epigenomes.

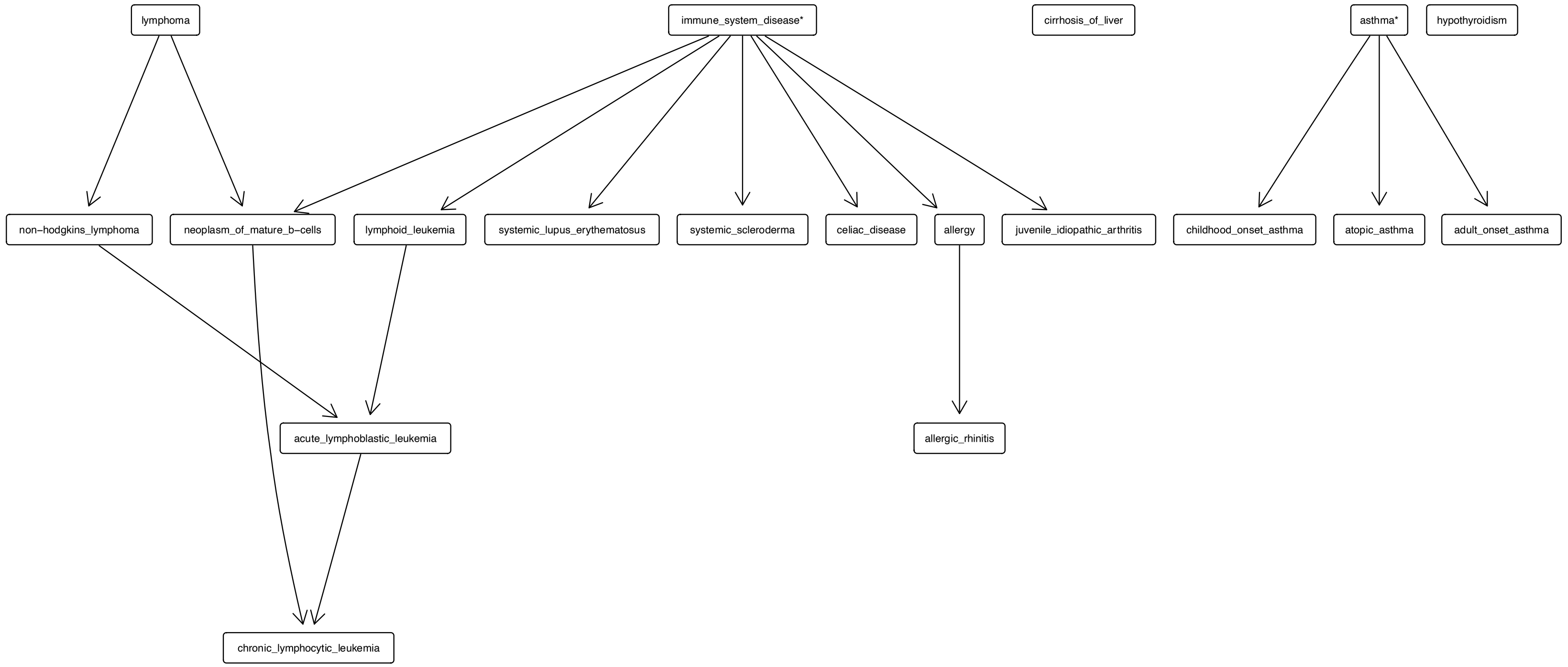


Supplemental Figure S10: ***Disease relationships for immune1 cluster***. The diseases placed at the top are more general than the diseases at the bottom. Arrow points from a more general term to a more specific term. A disease marked with one star indicates that it is not in this cluster but among the 111 diseases we studied. Diseases with two stars indicate that they are not among the 111 diseases.

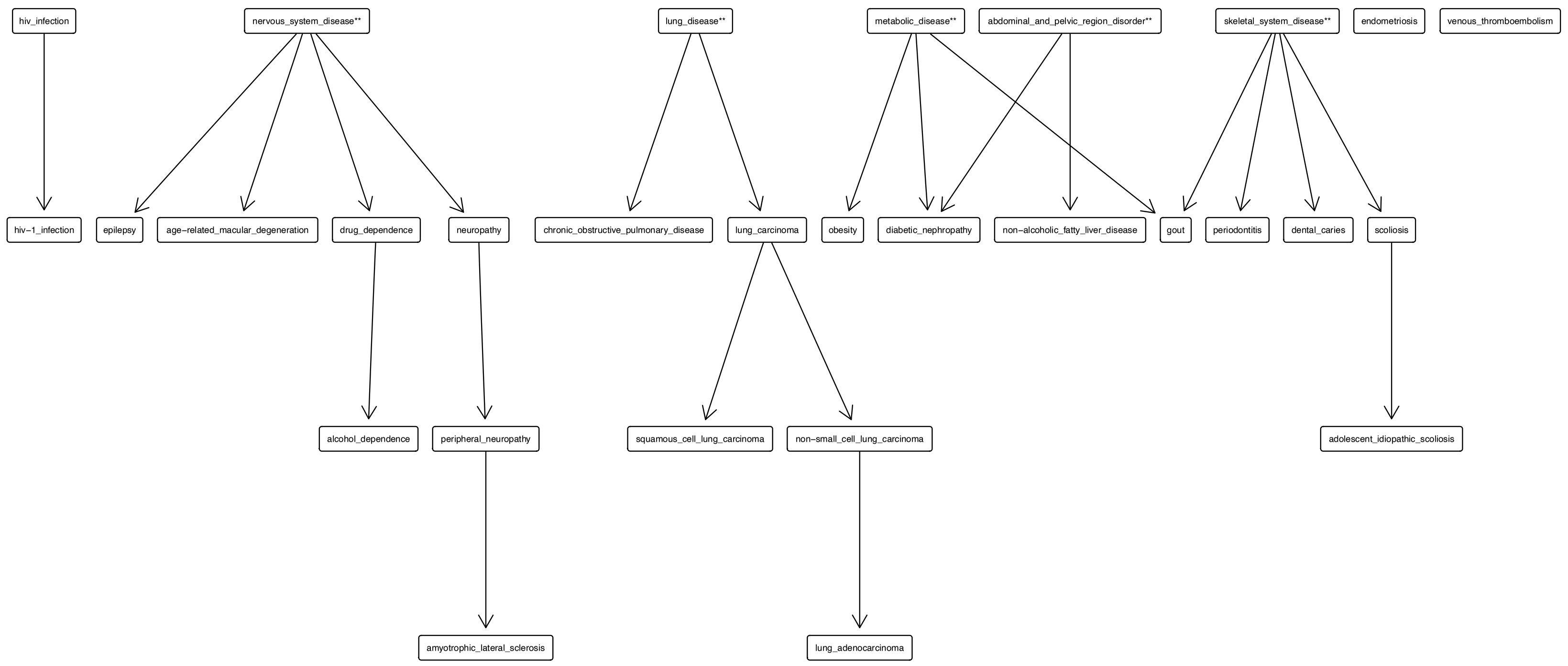


Supplemental Figure S11: ***Disease relationships for others cluster***. The diseases placed at the top are more general than the diseases at the bottom. Arrow points from a more general term to a more specific term. A disease marked with one star indicates that it is not in this cluster but among the 111 diseases we studied. Diseases with two stars indicate that they are not among the 111 diseases.

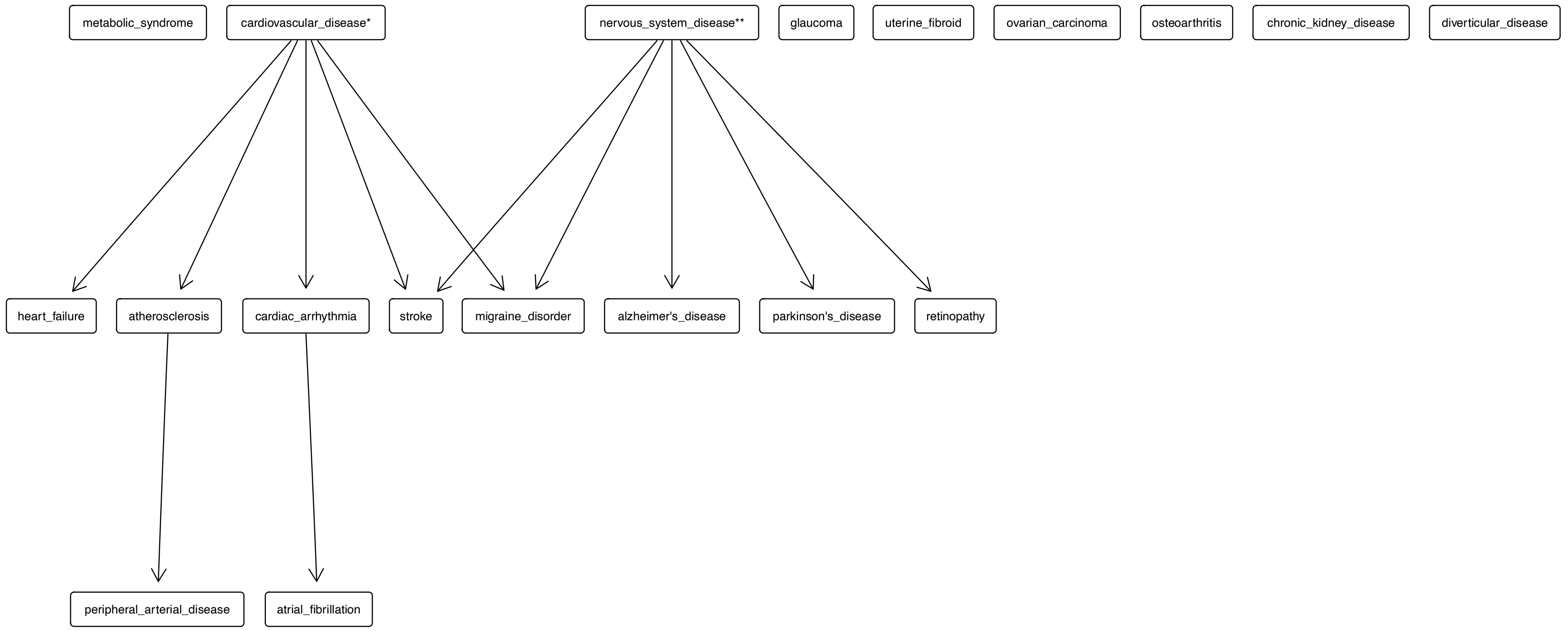


Supplemental Figure S12: ***Disease relationships for cardiovasular disease and others cluster***. The diseases placed at the top are more general than the diseases at the bottom. Arrow points from a more general term to a more specific term. A disease marked with one star indicates that it is not in this cluster but among the 111 diseases we studied. Diseases with two stars indicate that they are not among the 111 diseases.

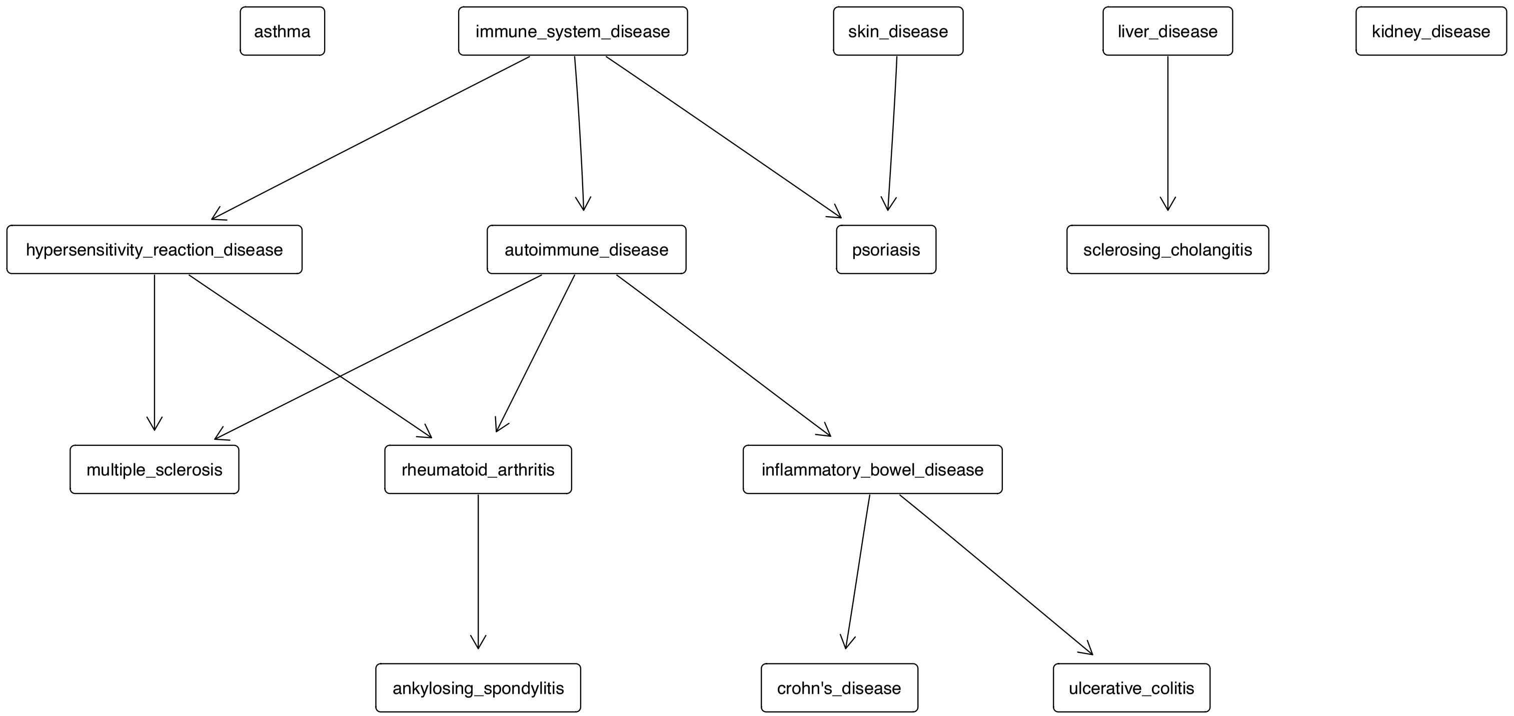


Supplemental Figure S13: ***Disease relationships for immune2 cluster***. The diseases placed at the top are more general than the diseases at the bottom. Arrow points from a more general term to a more specific term. A disease marked with one star indicates that it is not in this cluster but among the 111 diseases we studied. Diseases with two stars indicate that they are not among the 111 diseases.

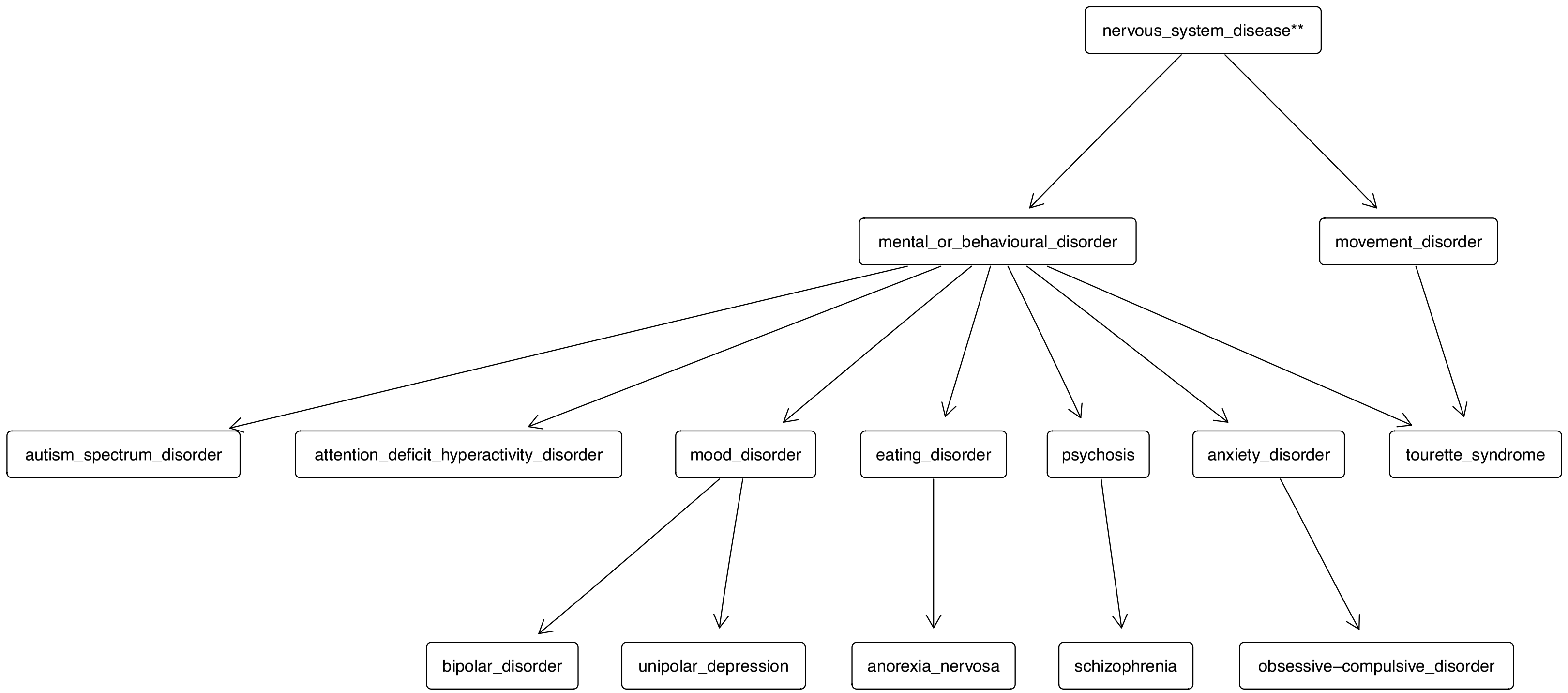


Supplemental Figure S14: ***Disease relationships for mental or behavioural disorder cluster***. The diseases placed at the top are more general than the diseases at the bottom. Arrow points from a more general term to a more specific term. A disease marked with one star indicates that it is not in this cluster but among the 111 diseases we studied. Diseases with two stars indicate that they are not among the 111 diseases.

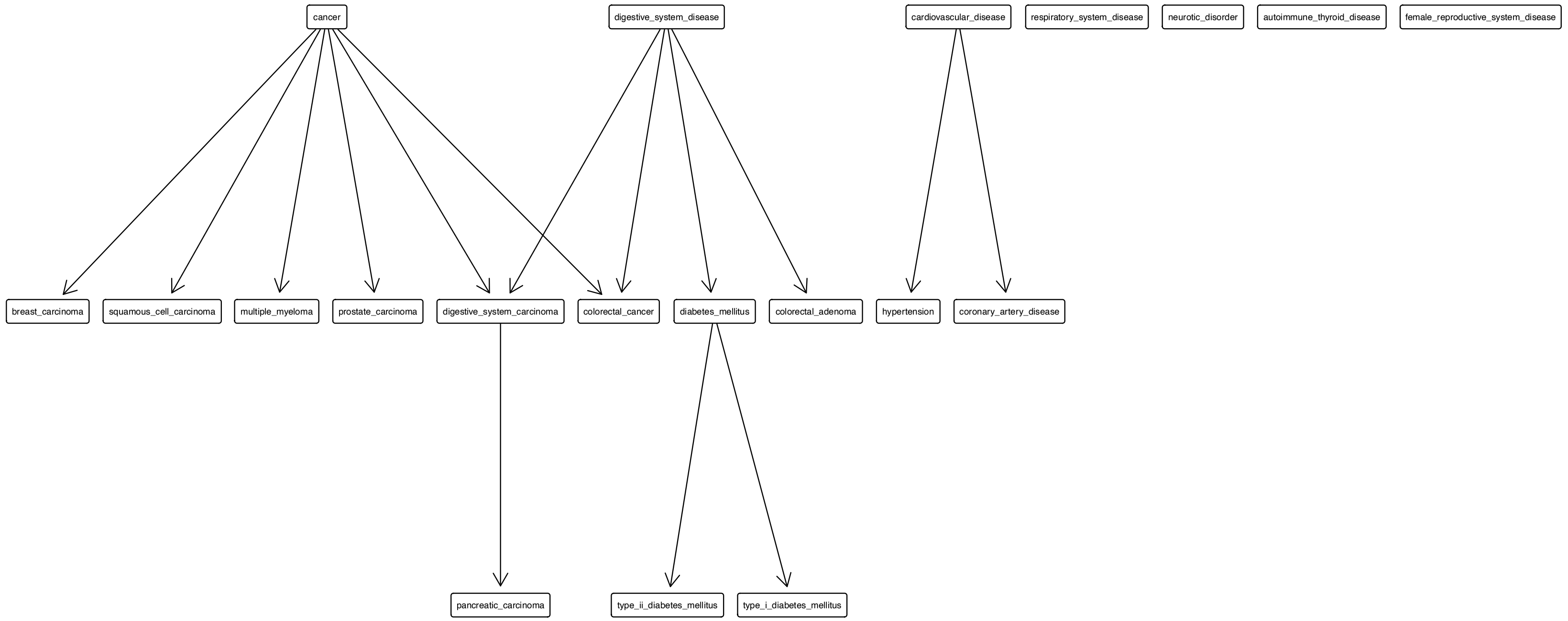


Supplemental Figure S15: ***Disease relationships for digestive and cancer cluster***. The diseases placed at the top are more general than the diseases at the bottom. Arrow points from a more general term to a more specific term. A disease marked with one star indicates that it is not in this cluster but among the 111 diseases we studied. Diseases with two stars indicate that they are not among the 111 diseases.

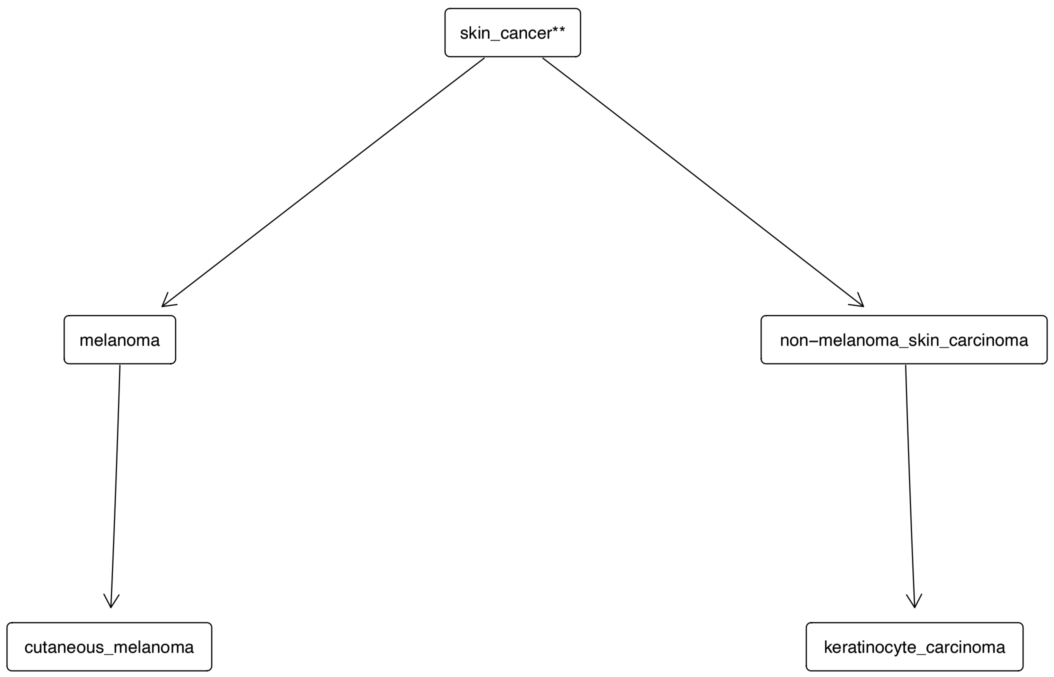


Supplemental Figure S16: ***Disease relationships for skin cancer cluster***. The diseases placed at the top are more general than the diseases at the bottom. Arrow points from a more general term to a more specific term. A disease marked with one star indicates that it is not in this cluster but among the 111 diseases we studied. Diseases with two stars indicate that they are not among the 111 diseases.

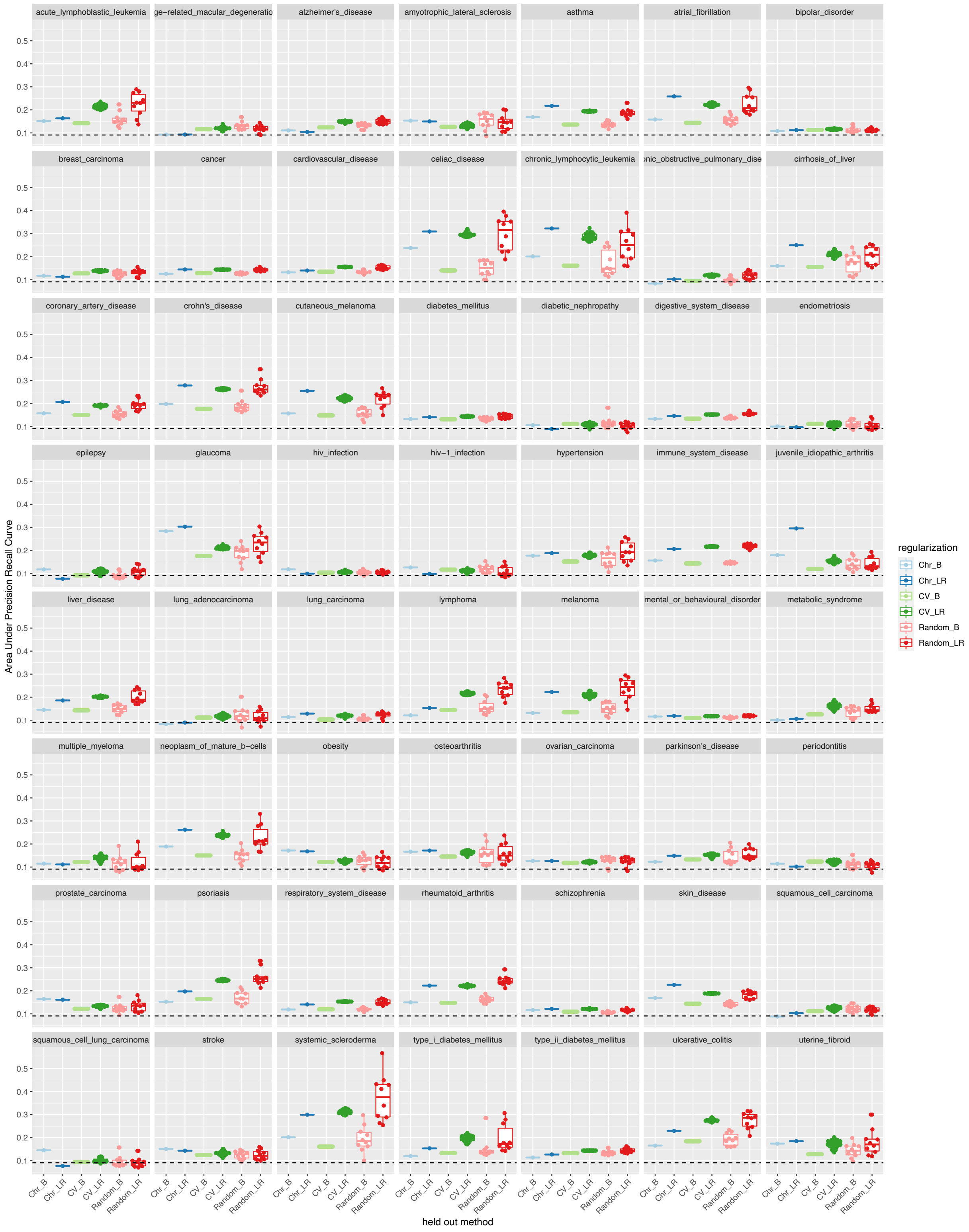


Supplemental Figure S17: ***Performance of Tissue-weighted (DHS) in different held-out strategies***. Chr-B: test set held out by chromosome, baseline; Chr-LR: test set held out by chromosome, logistic regression; CV-B: cross-validation, baseline; CV-LR: cross-validation logistic regression; random-B: randomly sampled test set, baseline; random-LR: randomly sample test set, logistic regression.

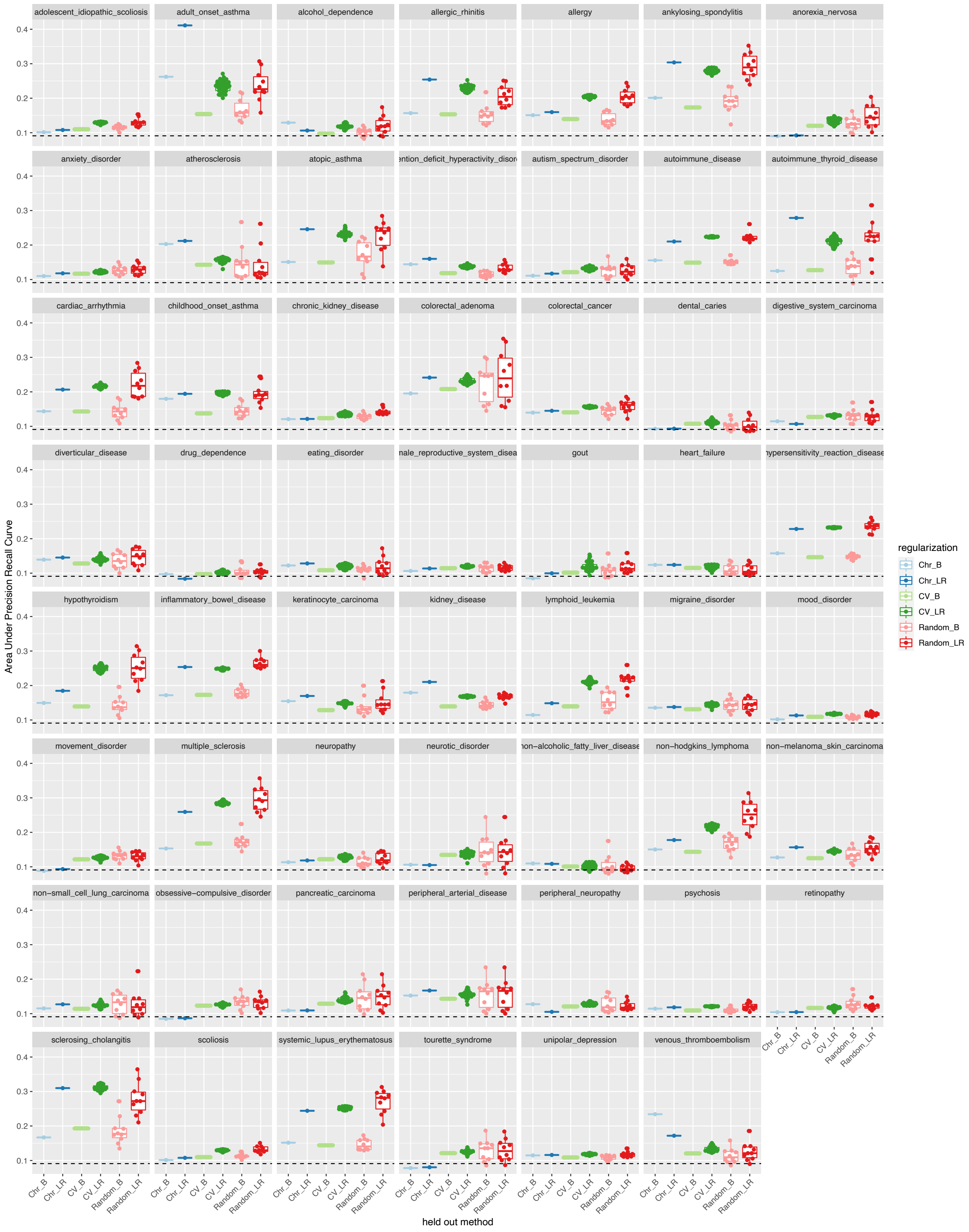


Supplemental Figure S18: ***Performance of Tissue-weighted (DHS) in different held-out strategies, continued***. Chr-B: test set held out by chromosome, baseline; Chr-LR: test set held out by chromosome, logistic regression; CV-B: cross-validation, baseline; CV-LR: cross-validation logistic regression; random-B: randomly sampled test set, baseline; random-LR: randomly sample test set, logistic regression.

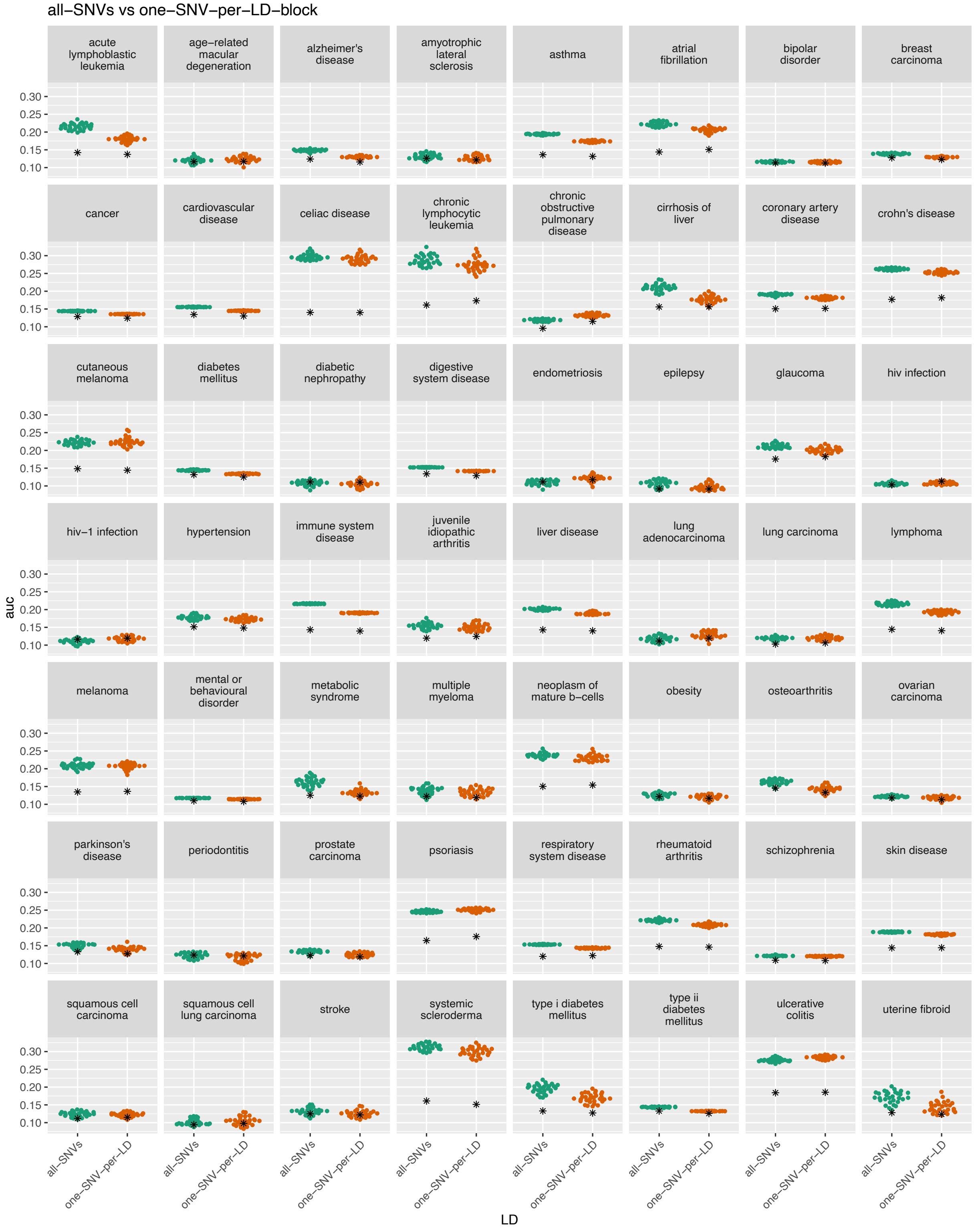


Supplemental Figure S19: ***Performance of Tissue-weighted (DHS) in all SNVs or representative SNVs (one SNV per LD block)***. Colored dots represent the performance of tissue-weighted (DHS) in all SNVs or representative SNVs. Stars represent the baseline performance (tissue-mean DHS) in all SNVs or representative SNVs.

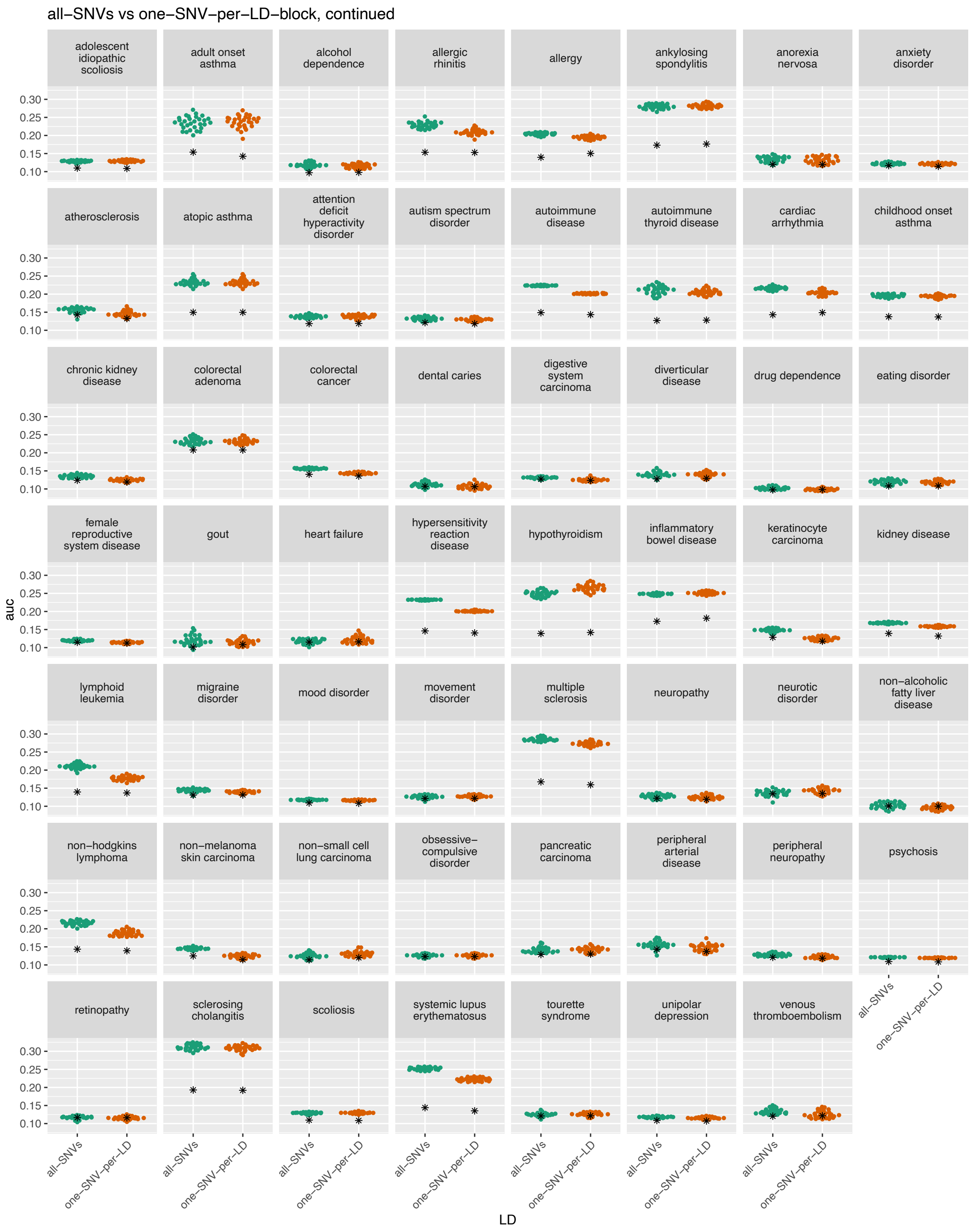


Supplemental Figure S20: ***Performance of Tissue-weighted (DHS) in all SNVs or representative SNVs (one SNV per LD block), continued***. Colored dots represent the performance of tissue-weighted (DHS) in all SNVs or representative SNVs. Stars represent the baseline performance (tissue-mean DHS) in all SNVs or representative SNVs.
